# Flexible and Highly-Efficient Feature Perception for Molecular Traits Prediction via Self-interactive Deep Learning

**DOI:** 10.1101/2023.07.30.23293391

**Authors:** Yang Hu, Korsuk Sirinukunwattana, Bin Li, Kezia Gaitskell, Willem Bonnaffé, Marta Wojciechowska, Ruby Wood, Nasullah Khalid Alham, Stefano Malacrino, Dan Woodcock, Clare Verrill, Ahmed Ahmed, Jens Rittscher

**Affiliations:** Nuffield Department of Medicine, University of Oxford, Oxford, UK; Department of Engineering Science, University of Oxford, Oxford, UK; Big Data Institute, Li Ka Shing Centre for Health Information and Discovery, University of Oxford, Oxford, UK; Nuffield Division of Clinical Laboratory Sciences, Radcliffe Department of Medicine, University of Oxford, Oxford, UK; Nuffield Department of Surgical Sciences, University of Oxford, Oxford, UK; Department of Cellular Pathology, Oxford University Hospitals NHS Foundation Trust, Oxford, UK; MRC Weatherall Institute of Molecular Medicine, University of Oxford, Oxford, UK; Nuffield Department of Women’s and Reproductive Health, University of Oxford, Oxford, UK; Ludwig Institute for Cancer Research, Nuffield Department of Clinical Medicine, University of Oxford, Oxford, UK; Oxford National Institute for Health Research (NIHR) Biomedical Research Centre, Oxford, UK

## Abstract

Predicting disease-related molecular traits from histomorphology brings great opportunities for precision medicine. Despite the rich information present in histopathological images, extracting fine-grained molecular features from standard whole slide images (WSI) is non-trivial. The task is further complicated by the lack of annotations for subtyping and contextual histomorphological features that might span multiple scales. This work proposes a novel multiple-instance learning (MIL) framework capable of WSI-based cancer morpho-molecular subtyping across scales. Our method, debuting as Inter-MIL, follows a weakly-supervised scheme. It enables the training of the patch-level encoder for WSI in a task-aware optimisation procedure, a step normally improbable in most existing MIL-based WSI analysis frameworks. We demonstrate that optimising the patch-level encoder is crucial to achieving high-quality fine-grained and tissue-level subtyping results and offers a significant improvement over task-agnostic encoders. Our approach deploys a pseudo-label propagation strategy to update the patch encoder iteratively, allowing discriminative subtype features to be learned. This mechanism also empowers extracting fine-grained attention within image tiles (the small patches), a task largely ignored in most existing weakly supervised-based frameworks. With Inter-MIL, we carried out four challenging cancer molecular subtyping tasks in the context of ovarian, colorectal, lung, and breast cancer. Extensive evaluation results show that Inter-MIL is a robust framework for cancer morpho-molecular subtyping with superior performance compared to several recently proposed methods, even in data-limited scenarios where the number of available training slides is less than 100. The iterative optimisation mechanism of Inter-MIL significantly improves the quality of the image features learned by the patch embedded and generally directs the attention map to areas that better align with experts’ interpretation, leading to the identification of more reliable histopathology biomarkers.

## 1 Introduction

Recent advances in computer vision and artificial intelligence (AI) have dramatically reformed computational pathology. With powerful deep neural networks (DNNs), whole slide image (WSI)-based morphological subtyping has become an emerging tool with great potential for future use in the clinic and in drug discovery. The core concept of morpho-molecular subtyping is to infer biologically relevant molecular traits directly from the morphological features presented in hematoxylin and eosin (H&E) histopathological samples, thus circumventing the need for expensive and time-consuming molecular assays^1^. The successful deployment of such methods can have a profound impact on cancer treatment, offering a cost-effective solution for personalized medicine that leverages the richness of the information contained in WSIs. Such techniques promise to alleviate the dependency on time-consuming and potentially costly gene sequencing^2,3^, enabling a quicker initiation of treatment for patients with distinct cancer biomarkers. However, morpho-molecular subtyping doesn’t aim to replace comprehensive genetic testing, but rather to supplement it. By bridging the gap between traditional histopathology and molecular profiling, we can potentially streamline the diagnostic process, making subtype-specific interventions more readily available. This advancement will bring us one step closer to the reality of cost-effective precision medicine.

Recent years have witnessed rapid adoption of whole slide imaging in both the clinic and research as the digitisation of physical histology samples enables automatic software analysis, advanced data management, and remote image viewing and conferencing. A large volume of recent research has been focused on analysing WSIs using DNNs. Various methodologies for improving diagnostic accuracy, prognostication, and identifying ambiguous and high-risk cases prioritised for detailed molecular testing and immunohistochemistry have been proposed^4–8^. However, applying DNNs such as convolutional neural networks (CNNs) directly on whole WSIs at full magnification can be very challenging due to the high spatial resolution of WSIs. The memory requirements for processing an entire WSI using a CNN in a single iteration are beyond the limits of current standard graphics processing units (GPUs). Hence, the commonly applied strategies for WSI processing usually involve splitting the image into small tiles and/or pre-compressing the image tiles into feature vectors to reduce the size of the input^9–16^. Another obstacle in applying deep learning on WSIs is the difficulty in obtaining reliable annotations. Informative disease-relevant histomorphological features can be rare and subtle occurrences. Extensive tile-level annotations may incur extremely high labour costs and are often impractical to acquire^17,18^. On the other hand, weakly-supervised learning-based methods model the tile-to-slide correlations to achieve slide-level predictions without the need for the presence of tile-level annotations at training. Because of this, weakly-supervised methods have garnered considerable attention for their adeptness in addressing the particular challenges inherent to the analysis of WSIs.

One family of weakly supervised learning models frequently used for classifying unannotated WSIs is multiple instance learning (MIL). Here, tiles cropped from a WSI are considered individual instances. The WSI is considered to be a bag containing these instances^20^. The original formulation of MIL for binary classification deploys a basic independent and identically distributed (i.i.d.) assumption for the instances, meaning that the label of each instance is assumed to be independently drawn from a Bernoulli distribution. The bag label is then conditioned on all the instance labels, and it is positive if a least one instance label is positive and negative if all instance labels are negative. Recent efforts have also included modelling the correlation between the instances, using architectures such as graph models or self-attention^21–23^.

Typical MIL methods used for analysing WSIs can be roughly separated into two categories. Instance-based methods optimise the subtyping likelihood of a subset of tile images that are representative of the whole slide^9,24^. In these methods, the randomness in the initial selection of representative tiles may lead to difficulties in optimisation convergence. Additionally, it relies on assigning pseudo-labels to representative tile-level features for slide-level modelling, which could inadvertently ignore the global morphological features. Another category of more complex methods is embedding-based, where the learning is generally separated into two stages: 1) Encoding: embedding tiles into an abstract feature space and 2) Aggregating: summarizing the tile embeddings for a slide-level embedding and then scoring the slide-level embedding. The goal of the encoding phase is to obtain compressed representations of tiles, which is usually performed using a pretrained neural network. The pretraining tasks can be ImageNet classification^25^, self-supervised learning^26,27^, or an easier task related to the target task^14^. The aggregator fuses the tiles embeddings of a WSI to produce a global representation and performs the final prediction. Due to the fact that informative regions may only occupy a small portion of the whole slide, attention-based pooling is often used to select potentially representative tiles while suppressing the contribution of other, noisy regions^20,28^. The attention-based pooling scheme is also frequently used in conjunction with multi-resolution representations, clustering, self-attention layers (e.g. Transformers), and graph-based models, enabling the model to integrate contextual information or prior knowledge regarding tissue morphology^11–13,29–32^. However, the majority of these methods do not optimise the tile-level encoder with respect to the prediction task in a closed-loop and instead resolve to pretrained encoders obtained from a proxy task agnostic to the downstream prediction. This approach limits the aggregator’s ability to perceive fine-grained information^33^. Consequently, most histopathological image analysis frameworks utilize large cohorts with hundreds or thousands of WSIs for training, compensating for sub-optimal fine-grained tile-level features^9,12,13,15,24,28^.

Predicting molecular traits from histomorphology poses some specific challenges that are less relevant in other computational pathology tasks for example tumour detection. Firstly, collecting gene sequencing-supported subtype annotations requires strict quality control, is expensive, and often, different molecular subtypes can exhibit visually similar phenotypes on H&E slides, making molecular subtypes not as distinguishable as other histopathological classification tasks^1^. As a result, successful recent research for molecular subtyping normally requires a considerable amount of training samples^1,34,35^. We typically leverage multi-omics data^36,37^, immunohistochemistry (IHC) stained images targeting proteins associated with bespoke phenotypes^32,38,39^, or manual pixel-level region of interest (ROI) annotations^39^ for training.

Given the cost and effort necessary to provide such additional information, well-annotated patient cohorts that are suitable for developing models for molecular subtyping are often small. Although they typically do not provide many patient samples, the tile-level statistics can still be abundant. The availability of a massive quantity of tiles may enable the exploration of detailed and extensive statistical analysis at a more granular level. In addition, representative features in highly heterogeneous disease contexts can appear in different tissue scales ranging from the cellular level to tissue level^40^. Pathologists usually cannot provide exact histopathological descriptions for specific molecular subtypes across scales. In many cases, whether the subtype-related image features are on the tissue or cellular level is practically unknown. In this paper, we present a novel MIL-based approach for WSI-based morpho-molecular subtyping. Unlike most existing frameworks that utilize pretrained tile-level encoders, our work, named Inter-MIL, enables end-to-end training of the tile-level encoder jointly with the aggregator, allowing more task-specific discriminative features to be learned at the tile level. To optimise the tile-level encoder, we employ a pseudo label propagating strategy which captures the interaction between tiles and slide-level labels. Notably, this strategy also improves the aggregator responsible for summarizing the tile-level features, leading to better global tissue features. Visualization reveals that during the iterative optimisation, the aggregator’s attention is generally directed to better align with regions that experts find significant. At the same time, the tile-level image features become more discriminative. Moreover, using gradient-based methods to examine encoder activations, we extract finer-grained attentions that capture cellular features within individual tiles, surpassing the capabilities of a task-agnostic pretrained encoder. With the proposed Inter-MIL, the tile-level features are extracted in a task-relevance fashion. These cellular level attentions also demonstrate concordance with pathologists’ interpretations, indicating that capturing nuclei and other fine-grained features of tiles^41,42^ strongly associated with the task is beneficial for delivering quantitative features of cytopathology^43,44^ to the MIL framework. Moreover, these biologically relevant features hold the promise of unlocking the discovery of novel microanatomical structures in a data-driven fashion enabled by AI models.

The main methodological contributions and insights obtained by this study are summarised with the help of Figure 1: **(a)** <u>Diverse set of molecular subtyping tasks</u> - We consider four very different molecular subtyping tasks, including the prediction of high Epithelial-mesenchymal transition (EMT) in serous epithelial ovarian cancer (SOC)^45,46^, the prediction of Kirsten rat sarcoma viral oncogene (KRAS) mutation status in colon^47^ and lung cancer^24,48^, epidermal growth factor receptor (EGFR) mutation status in lung cancer^24,49^, and Human epidermal growth factor receptor 2 (HER2) amplification in breast invasive cancer^50^. **(b)** <u>Novel Inter-MIL framework</u> - We introduce an iterative optimisation through communication between features with local- and large-scale granularity. Inter-MIL improves the learning efficiency of MIL on small histopathological datasets. The proposed Inter-MIL introduces optimising steps at various feature scales for both the tile-level encoder and slide-level aggregator. **(c)** <u>More representative features</u> - Inter-MIL searches for representative features of molecular subtypes from multiple scales, allowing the identification of cytopathological features even within the tiles, and improving the search for coarse-grained features. **(d)** <u>Inter-MIL features improve discrimination</u> - Inter-MIL reshapes the tile-level feature space, making the visual features of different molecular subtypes more distinguishable, thereby reducing the difficulty of subtyping new samples.

**Figure 1.**
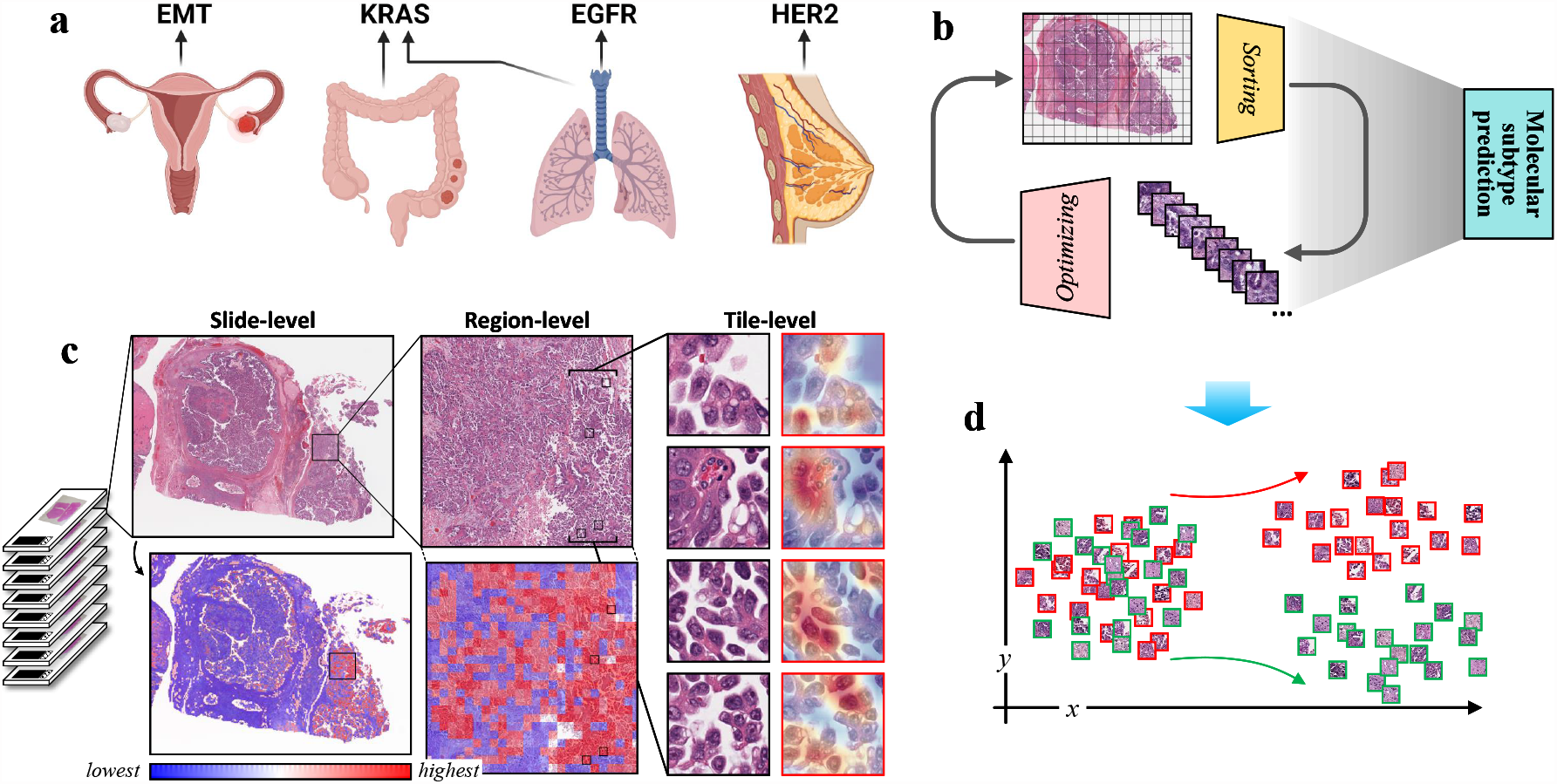
Molecular trait prediction and feature investigation: task, method, and vision. **a** Task: Prediction of 4 molecular traits on datasets of 4 cancer types. **b**, Method: Our novel Inter-MIL approach to drive self-interaction between global biopsy WSI features and fine-grained tile-level features. **c**, Vision: From left to right, presented at slide-level, region-level, and fine-grained tile-level attention interpretation of models, where the Grad-CAM tool^19^ provides the detail-to-nucleus attention interpretation of tiles. **d**, Vision: The proposed Inter-MIL approach is expected to provide a more discriminative feature space for informative tiles from all slides.

## 2 Results

### Datasets and model variants

The datasets used to validate the proposed approach and explain the nomenclature of variants of the proposed method are being presented. The experiments are conducted on four subtyping tasks: 1) ***OV-EMT:*** Approximately 20% of serous ovarian cancers (SOCs) are classified as Epithelial-to-mesenchymal transition–high (EMT-high) tumours, which are associated with poor survival^46^. Here, we analysed 70 WSIs from TCGA-OV dataset with a binary EMT status (38 EMT-high vs. 32 EMT-low); 2) ***COLU-KRAS:*** Mutations in the Kirsten rat sarcoma viral oncogene (KRAS) gene are often associated with different cancer types, including lung cancer, colorectal cancer^47,48^. The presence of KRAS mutations in colorectal cancer can have implications for treatment decisions^52^. Here we present a combined cohort of 112 WSIs with KRAS mutation status (44 mutated vs. 68 wild-type) from TCGA-COAD and TCGA-LUAD datasets; 3) ***LU-EGFR:*** Detection of Epidermal Growth Factor Receptor (EGFR) mutations is now a standard part of the diagnostic workup for patients with non-small cell lung cancer (NSCLC), as it helps guide treatment decisions^53^. Here we utilized 261 WSIs from TCGA-LUAD dataset for subtyping EGFR mutation status (75 mutated vs. 186 wild-type); 4) ***BR-HER2:*** Human Epidermal Growth Factor Receptor 2 (HER2) is a protein that is overexpressed in approximately 15-20% of breast cancers. HER2-positive breast cancers tend to be more aggressive and less responsive to hormone treatments compared to HER2-negative cancers^54^. 415 WSIs from the TCGA-BRCA dataset where HER2 status was determined based on fluorescence amplification in situ hybridization (FISH) expression (77 positives vs. 338 negatives) were used. For annotations, EMT status used in *OV-EMT* is available in^45,46^ while the subtype labels for the rest of the tasks are available in the TCGA data repository^55^.

In this paper, a self-interactive multi-instance learning (Inter-MIL) approach is proposed, which utilizes two modules to model fine-grained tile-level features and global slide-level features, respectively. These two modules interact with each other to achieve mutual optimisation. For Inter-MIL, we define the following variants: 1. Inter-MIL-b, which is a simplified version without random tiles-level features; 2. adInter-MIL, which is based on the standard Inter-MIL but adds adversarial training^56^ on noisy tiles. In addition to these variants, we also design a pre-training module for aggregation classifiers in MIL. This is a general hot-swappable module suitable for most MIL methods, so we use the prefix “PT-” to indicate that this module is applied. More algorithm and technical details can be found in the Method section (4).

### Evaluation on slide-level subtyping

The performance of Inter-MIL is evaluated in terms of 1) the average and variance of the area under the receiver operating characteristic curve (AUC), and 2) the balanced accuracy (BACC) across multiple test folds is discussed.

As shown in Tables 1 and 2, the Inter-MIL variants, compared to the baseline Gated-AttPool method, achieved at least a 6% improvement and demonstrate the best performance. As illustrated in Figure 2-a, Inter-MIL and its variants outperform the baseline method on all four tasks. Investigating the optimisation process, as shown in Figure 2-c, the loss of Inter-MIL in the early iterations is on par with the baseline method Gated-AttPool, but then the training rapidly converges after the interactive optimisation of tile-level encoder. Figure S-7-a shows that Inter-MIL and adInter-MIL have similar loss-declining training logs.

**Table 1.** Results on multiple molecular subtyping tasks with ROC-AUC *±* variance (%) over 10 runs for task *OV-EMT* and 5 runs for tasks *COLU-KRAS, LU-EGFR*, and *BR-HRER2*.

| Methods | *OV-EMT* | *COLU-KRAS* | *LU-EGFR* | *BR-HER2* |
| --- | --- | --- | --- | --- |
| CNN-MIL<sup>9</sup> | 59.86 ± 1.11 | 50.56 ± 0.15 | – | – |
| AttPool<sup>20</sup> | 61.38 ± 1.35 | 61.41 ± 0.78 | – | – |
| Gated-AttPool<sup>20,28</sup> | 62.46 ± 1.13 | 59.94 ± 0.06 | 64.15 ± 0.39 | 54.84 ± 0.14 |
| CLAM<sup>13</sup> | 62.62 ± 1.10 | 62.18 ± 0.80 | 65.17 ± 0.25 | 54.17 ± 0.11 |
| FocAtt-MIL<sup>14</sup> | 56.89 ± 1.86 | 63.76 ± 1.05 | 64.19 ± 0.12 | 53.53 ± 0.15 |
| Inter-MIL (ours) | 71.91 ± 0.50 | 64.78 ± 0.18 | 69.81 ± 0.12 | 63.08 ± 0.04 |
| Inter-MIL-b (ours) | 68.53 ± 0.21 | 64.01 ± 0.3 | 67.99 ± 0.02 | 62.00 ± 0.07 |
| adInter-MIL (ours) | **74.55 ± 0.43** | **66.34 ± 0.31** | **70.65 ± 0.08** | **63.21 ± 0.03** |
| *PT - Gated-AttPool (ours)* | 64.18 ± 1.01 | 62.34 ± 0.70 | 65.21 ± 0.25 | 57.21 ± 0.22 |
| *PT - Inter-MIL (ours)* | 74.41 ± 0.26 | 70.38 ± 0.28 | 70.15 ± 0.24 | 62.94 ± 0.03 |
| *PT - adInter-MIL (ours)* | 77.00 ± 0.40 | 71.38 ± 0.11 | 71.33 ± 0.08 | 64.02 ± 0.11 |

**Table 2.** Results on multiple molecular subtyping tasks with BACC *±* variance (%) over 10 runs for task *OV-EMT* and 5 runs for tasks *COLU-KRAS, LU-EGFR*, and *BR-HRER2*.

| Methods | *OV-EMT* | *COLU-KRAS* | *LU-EGFR* | *BR-HER2* |
| --- | --- | --- | --- | --- |
| Gated-AttPool<sup>20,28</sup> | 70.45 ± 1.87 | 59.41 ± 0.08 | 60.34 ± 0.20 | 53.29 ± 0.02 |
| CLAM<sup>13</sup> | 69.00 ± 1.08 | 62.97 ± 0.27 | 61.11 ± 0.20 | 55.63 ± 0.11 |
| FocAtt-MIL<sup>14</sup> | 61.71 ± 0.72 | 61.74 ± 0.10 | 57.32 ± 0.02 | 53.56 ± 0.02 |
| Inter-MIL (ours) | 84.86 ± 0.45 | 64.50 ± 0.45 | 66.04 ± 0.33 | 56.69 ± 0.07 |
| adInter-MIL (ours) | **85.45 ± 0.48** | **65.85 ± 0.24** | **69.56 ± 0.03** | **63.21 ± 0.03** |
| *PT - Gated-AttPool (ours)* | 71.45 ± 0.89 | 62.49 ± 0.36 | 61.19 ± 0.21 | 57.40 ± 0.14 |
| *PT - Inter-MIL (ours)* | 85.45 ± 0.38 | 67.86 ± 0.42 | 69.81 ± 0.17 | 58.52 ± 0.04 |
| *PT - adInter-MIL (ours)* | 85.77 ± 0.61 | 69.78 ± 0.16 | 70.87 ± 0.15 | 63.67 ± 0.14 |

**Figure 2.**
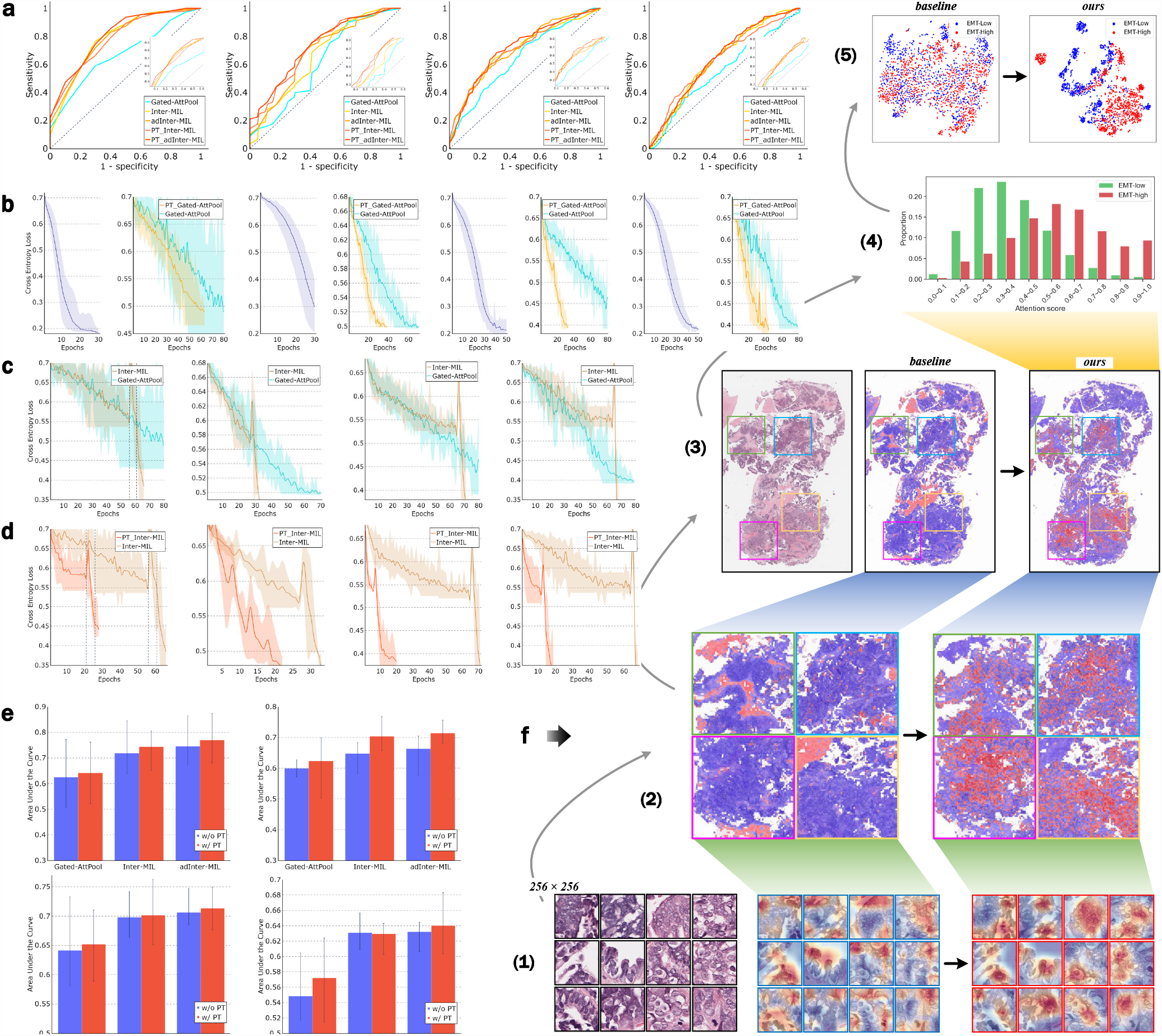
Overview of the results and the interpretations. **a** AUC-ROC curves of different models, for tasks: *OV-EMT, COLU-KRAS, LU-EGFR*, and *BR-HER2*, from left to right. Likewise below. **b**, log of loss for MIL aggregator pretraining and log of loss comparison for Gate-AttPool models with/without aggregator pretraining. **c**, log of loss comparison for Gate-AttPool models and Inter-MIL models. **d**, log of loss comparison for Inter-MIL models with/without aggregator pretraining. **e**, AUC performance comparison for Gate-AttPool, Inter-MIL, and adInter-MIL models with/without aggregator pretraining. From left to right in **a∼d** and left-top to right-bottom in **e**, the charts illustrate the results on *OV-EMT, COLU-KRAS, LU-EGFR*, and *BR-HER2* tasks. **f**, Model interpretation comparison at various scales of the baseline model (on the left) and adInter-MIL model (ours, on the right), which uses the case of *OV-EMT* task as an instance. From bottom to top, **f-(1)**, Fine-grained scale. Gradient activation heatmaps of the instanced tile images. **f-(2) and f-(3)**, Macroscopic scale. Attention heatmaps on representative regions, and their corresponding location on the slide. **f-(4)**, Attention score statistics in the slide-level. Attention score distribution of tiles with different prediction results on EMT-low/high. **f-(5)**, Feature space visualization at the test cohort level. The feature space t-SNE^51^ mapping of high informative tiles from all test slides.

In addition, pretraining the aggregation classifier accelerates the optimisation of both Gated-AttPool and Inter-MIL series. As evidenced by Figures 2-b, 2-d, and S-7-b, after the aggregation classifier was pretrained using contrastive learning, the convergence of the training is expedited by at least 10 epochs. This acceleration was even more pronounced in tasks such as *LU-EGFR* and *BR-HER2*, which have more training data, where the convergence was more than 40 epochs ahead. With the pre-training of aggregator for 30 epochs on tasks *OV-EMT* and *COLU-KRAS*, and for 40 epochs on the other two tasks with bigger datasets, the models continue to improve subtyping performance on tasks *OV-EMT* and *COLU-KRAS*, but not significantly on the tasks *LU-EGFR* and *BR-HER2*, as shown in Tables 1, 2, and Figure 2-e. In general, across the four subtyping tasks, Inter-MIL consistently demonstrated superior performance in terms of AUC and BACC, even with limited training data, and it enhanced the efficiency of MIL training. In addition, Figure S-7 provides the comparison results for Inter-MIL on different hyper-parameter settings, on tasks *OV-EMT* and *COLU-KRAS*, showing that there may be some fluctuations in performance on different hyperparameters, but it generally remains better than the baseline. More analysis and technical details can be found in the Discussion (3) and Method (4) sections.

### Interpreting the model at various scales

We highlight the interpretability of Inter-MIL under weakly supervised conditions, which provides insight into the optimisation process of features at various scales, from fine-grained tile-level features to the distribution of global biologically relevant instances. Figure 2-f showcases a test case from the *OV-EMT* task, which demonstrates the interpretation workflow from bottom to top: (1) Inter-MIL iteratively optimises the encoder for tile instances. With different models, we use the Gradient-weighted Class Activation Mapping tool (Grad-CAM) to generate the gradient-based attention heatmaps within individual tiles. Comparing the attention regions on example tiles between the baseline method and Inter-MIL, we observe that the high attention regions move toward cell nuclei areas in Inter-MIL as the tile level-encoder is iteratively optimised. (2) and (3) Macroscopically, we observe that the attention map also shifts from non-cancerous regions to tumour regions at the regional/global level during the optimisation. (4) We analyse the attention value distribution statistics on the exemplar slide and find that tiles with the EMT prediction score ⩾ 0.5 receive higher attention in EMT-high cases while the EMT prediction score *<* 0.5 co-occurs with higher attention in EMT-low cases. (5) By visualizing the feature space of representative tiles in all test slides, we find that the tile-level features learned by Inter-MIL are significantly more differentiable than the baseline for tiles of different subtypes (i.e., EMT-Low vs EMT-high). Therefore, the evolution of features and attention maps, the consistency of model attention and predictions, and an analysis of the latent feature space are now illustrated on a set of concrete examples.

### Evolution of tile-level attention

Inter-MIL iteratively optimises the **tile-level** encoder to model fine-grained vision features, learning increasingly detailed histological features to enable MIL’s instance-bag aggregator to better assess the representativeness of each tile for subtyping. Figure 3 illustrates the evolution of the attention distribution on tiles within a slide as the adInter-MIL improves the feature representation. After multiple rounds of interactive training, the model’s attention shifts from the tiles of background tissue to the tiles on or near tumour regions (Fig. 3-a and b). Furthermore, after one interactive training, the attention distribution of tiles in different regions has changed substantially, while round-3 is further fine-tuned based on round-2. Figure 3-a shows a histology slide which contains a blood clot. After the third round of interactive training, the model no longer pays attention to this area which does not have any diagnostic relevance. Figure 3-b presents an example where the attention shifts away from stromal tissue, instead, the model focuses more on the tumour areas, even showing the fissures between thin-strip-like tumour areas.

**Figure 3.**
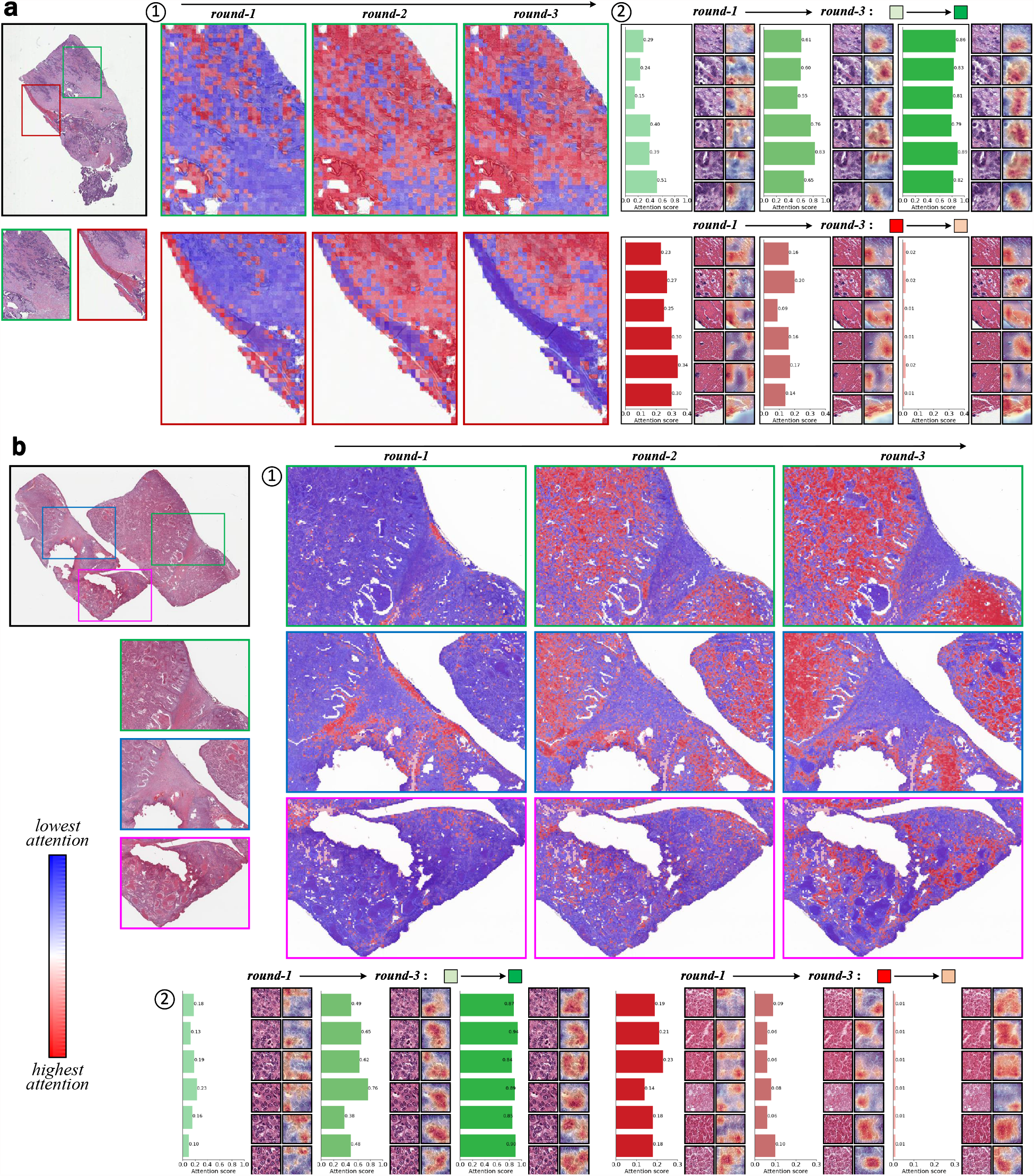
Attention evolution of tile-level features after each self-interaction round. **a** Example test case from the *OV-EMT* task. **b**, Example test case from the *COLU-KRAS* task. The images to the leftmost show the location of the example regions in the original WSIs. ➀ shows the evolution of the attention heatmap in different slide regions. Colour transition from blue to red indicates a rise in attention and vice versa. ➁ examples of tiles highly informative to the morphological classification task (green histograms) and of low relevance to the task (red histograms). The histogram, tile image and its attention heatmap demonstrate the attention evolution of these regions. Here in both presented scenarios, attention scores increase over time for tiles representing densely nuclear regions, and decrease for tiles containing connective tissue.

When contrasting tumour and non-tumour regions (see Figures 3-a and b), it can be observed that the tiles with gradually increasing attention mainly come from tumour regions, while the tiles with gradually decreasing attention come from background tissue regions such as stroma or blood. Especially in round-1, which has not yet undergone self-interactive training, the attention of connective tissue tiles is similar to the attention of tumour tiles or even higher, while after several rounds of self-interactive training, the attention of connective tissue tiles is reduced to zero. We further examine the fine-grained heatmaps within example tiles using gradient-based activation maps. The model’s attention within the tumour tiles gradually concentrates on regions containing nuclei from less informative scatterings. Even in non-tumour tiles containing a few cells, the model’s attention will be around them; otherwise, it maintains scattered vision heeding if there is no cell in the tile. This suggests that the tile-level encoder is trained to attend to entities that are potentially more informative.

Additional comparisons of attention maps of different methods can be found in Figures S-9, S-10, and S-11. More examples of the model’s attention-shifting evolution process can be found in Figure S-12. Figures S-13 and S-14 show the top attention tiles given by adInter-MIL and baseline methods. The fine-grained attention maps are improved and become concentrated around informative features such as nuclei after adInter-MIL interaction training, with the tile-level encoder being optimised. All these results show that the tile-level attention score assignment becomes more sensible as we iteratively optimise the tile-level encoder via self-interactive training to obtain more fine-grained features.

### Consistency across attention and different classes

In each optimisation round, Inter-MIL picks high-attention tiles, assigns them slide-level subtype labels, and trains the tile-level encoder. So, the other tiles are also able to obtain subtype classification scores after the optimisation of the tile-level encoder. In this section we investigate whether, following the interactive optimisation of Inter-MIL, the tiles from high-attention areas across different molecular subtypes simultaneously receive higher predictive scores aligned with their specific subtype from the classifier based on the tile-level encoder. For instance, for EMT-high slides, do the tiles in the high-attention areas also garner scores that lean more towards an EMT-high classification? Similarly, in EMT-low slides, do the tiles of high attention receive scores that favour an EMT-low classification? We demonstrate this by presenting positive and negative examples from tasks *OV-EMT* and *COLU-KRAS* in Figure 4. For each example, the left shows the original slide, the middle visualisation shows the attention value for each tile, and then the right shows the fine-grained classification score on each tile, in which the attention value is taken from the bag-of-tiles aggregator, and the fine-grained classifier has been iteratively trained on selected highly-informative tiles. We can see that for the EMT-low example, the classification outcome corresponding to the high attention area is closer to 0, while in the EMT-high example, the classification outcome corresponding to the high attention area is close to 1. A similar observation holds for the KRAS-no and KRAS-yes examples.

**Figure 4.**
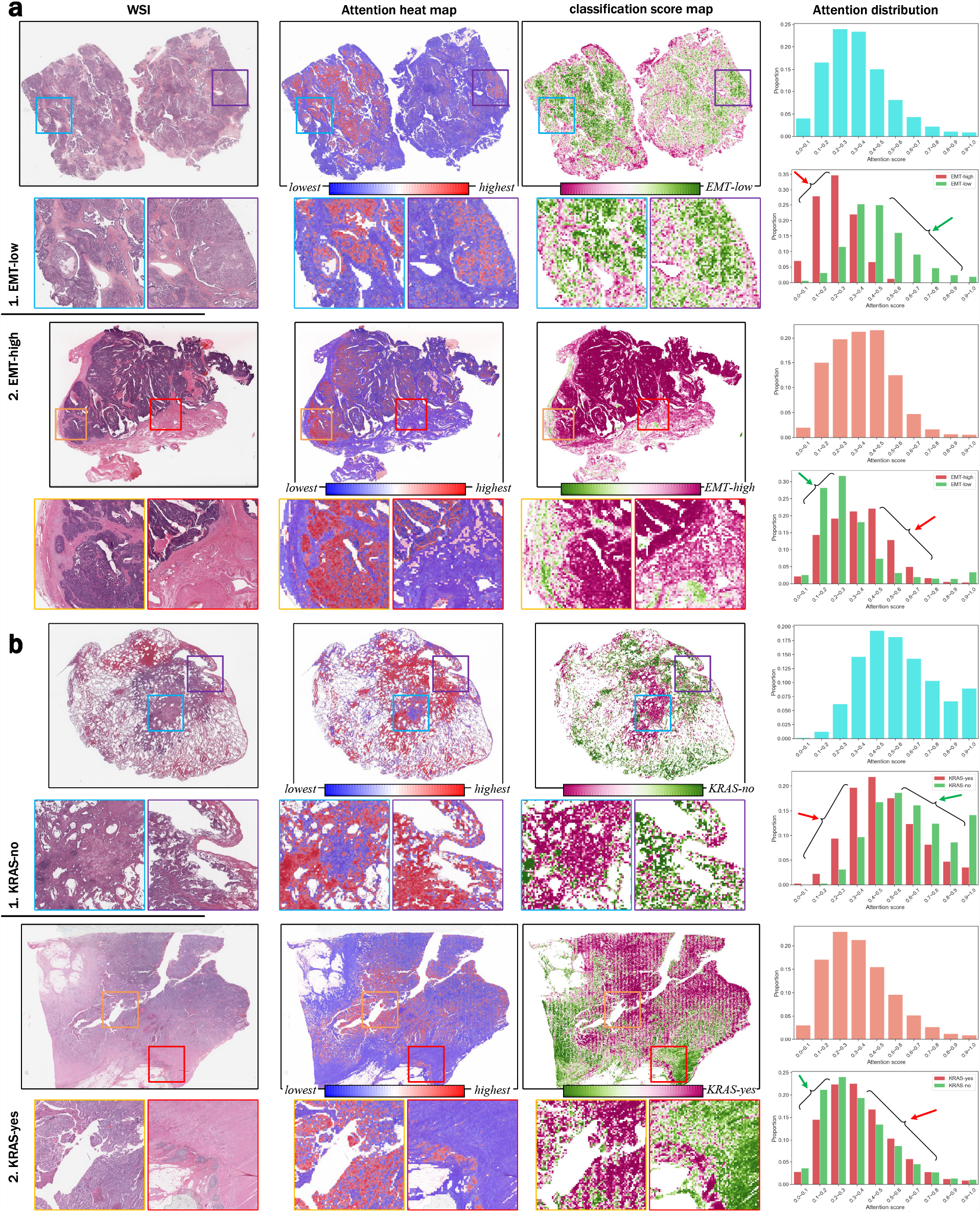
Distributions of slide-level attention and classification scores in two morpho-molecular subtyping classification tasks. **a** Example cases from the *OV-EMT* classification task, top: EMT-low case, bottom: EMT-high case. **b**, Example cases from the *COLU-KRAS* task, top: KRAS-no case, bottom: KRAS-yes case. For both **a and b**, from left to right: 1. the original WSI and the selected regions of interest; 2. attention heatmap; 3. classification score map; 4. Tile-attention histograms. Top: the proportion of tiles in the different attention ranges, bottom: proportions of tiles with prediction results of EMT-low/high (KRAS-no/yes). Here, we observe that in cases of different subtypes, tiles with higher attention obtain prediction scores that correspond more closely to their subtypes.

Additionally, the statistical analysis on the right of Figure 4 presents the attention distribution of the tiles on the example slides. The upper figure shows that the number of tiles in each attention range roughly follows a normal distribution. In the lower figure, the red bar represents the attention range proportion of tiles which are classified as EMT-high and KRAS-yes (by the tile-level classifier, with prediction score > 0.5), while the green bars show the proportion of tiles and these tiles’ classification results are EMT-low and KRAS-no (by the tile-level classifier, with prediction score < 0.5). In the cases of EMT-low and KRAS-no, we observe that the high attention tiles yield tile-level predictions closer to 0 (refers to EMT-low/KRAS-no class), whereas the high attention tiles in the EMT-high and KRAS-yes cases yield tile-level predictions closer to 1 (refers to EMT-high/KRAS-yes class).

### More discriminative features in latent space

In the previous section, we presented cases that demonstrate the agreement between the classification likelihood and attention distribution of tiles at the slide level. In this section, we demonstrate the proposed Inter-MIL leads to more discriminative histological features. 2D projections of the features obtained from the test set are used for illustration.

Figure 5-a displays the feature distribution of the 100 highest and lowest attention tiles of all slides in the *OV-EMT* test set. It is mapped to the 2-dimensional coordinates with help of the t-SNE^51^ dimensionality reduction method. Green and yellow dots refer to the lowest-attention tiles from EMT-low/high slides, respectively, while blue and red dots refer to the highest-attention tiles from EMT-low/high slides. The distribution of tile features with high attention is clearly different from that of low attention tiles, both in the baseline GatedAttPool-MIL model (denoted as model ‘X’ in the figure) and adInter-MIL model (denoted as model ‘Y’ in the figure). However, there is a noticeable difference between model GatedAttPool-MIL and model adInter-MIL as the distributions of high attention tiles of EMT-low and EMT-high are clearly distinguishable in adInter-MIL model, but not in model GatedAttPool-MIL. Representative tiles from high-attention EMT-low, high-attention EMT-high, and low-attention tiles are shown on the right side. It is noticeable that the low-attention tiles and high-attention tiles exhibit distinct visual features, while the differences among the high-attention tiles between the EMT-low and EMT-high subtypes are more subtle since the differentiation of the two subtypes might depend on fine-grained features in the nuclei level.

**Figure 5.**
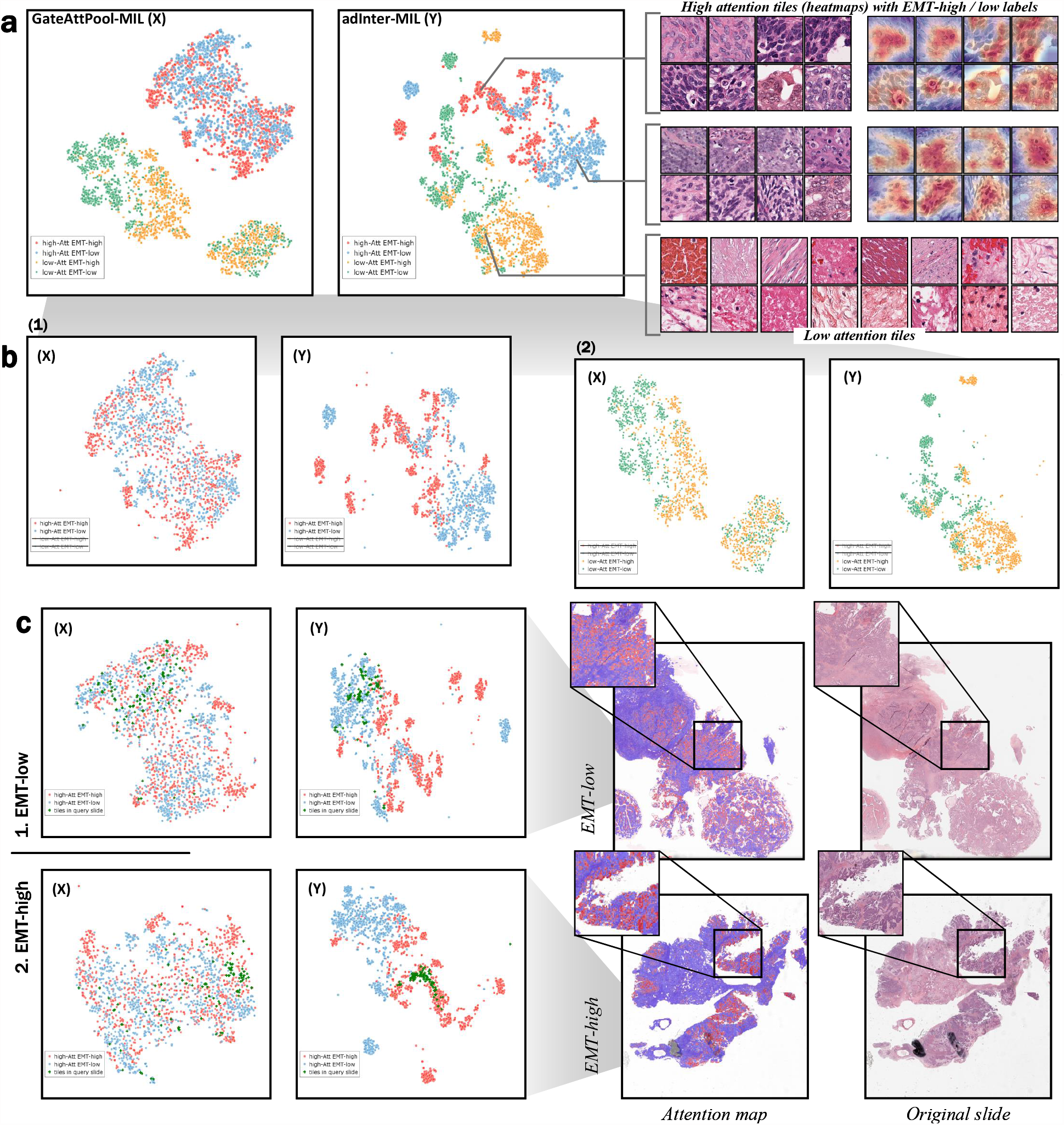
A comparison of the feature spaces of all the representative tiles in the *OV-EMT* dataset for two of trained models. **a** Left: the distribution of highly informative tiles and tiles with low task relevance in the learned cohort feature space, for the GatedAttPool-MIL (X) model and the adInter-MIL (Y) models; Right: example highly informative EMT-high/EMT-low tiles and examples of tiles without discriminative features. **b**, Comparison of the feature spaces of the (X) and (Y) models for highly informative tiles and tiles with low task relevance. **c**, The distributions of the highly informative tiles (green) taken from the two example cases: with EMT-low and EMT-high status respectively over the feature spaces of the two tested models. It can be seen that informative tiles form clearer, more separate clusters in the feature space of the adInter-MIL model. The tiles corresponding to the two example cases are located within the clusters corresponding to their correct label in the adInter-MIL feature space.

Figure 5-b illustrates the difference in feature distribution between model GatedAttPool-MIL and model adInter-MIL for high-attention and low-attention tiles, respectively. We note that the features of the high-attention tiles benefit more from our Inter-MIL approach and become more discriminating in the feature space, while the low-attention tile features are less affected. This suggests that the improvement in classification performance is more significant on highly informative features rather than low attention regions that are potentially noisy.

Figure 5-c further presents examples of querying high-attention tiles in the feature space. One example is EMT-high, and another is EMT-low. Dark green dots indicate high attention tiles from queried slides. We can observe that, whether it is EMT-low or EMT-high, the query points are closer to the respective subtype cluster and farther away from the other subtype clusters in the feature distribution of model Y’s. In contrast, querying different subtypes with model X is not as convenient (the green dots are more scattered in the latent space).

More results regarding the learned tile-level features can be found in Figures S-16 and S-17. Among them, Figure S-16 provides insights into the impact of sampling varying numbers of top attention tiles in all four molecular subtyping tasks. The results reveal that after self-interactive training, the feature spaces of different subtypes become more separated. However, it is worth noting that on the *BR-HER2* task, the distinguishability of the feature space generated by adInter-MIL is slightly lower compared to the other tasks, a portion of HER2-Neg and HER2-Pos tile-level features are still mixed together in the feature space. Moreover, in Figure S-17, we can find that for all four tasks, the features of query tiles from a specific slide are clustered with the tile features of the same subtype from other slides. More specifically, in the feature space provided by Inter-MIL and adInter-MIL, for a given test slide, the majority of its high-attention tiles’ neighbours come from high-attention tiles from slides of the same molecular subtype. In contrast, for GatedAttPool-MIL, a significant portion of the neighbours of its high-attention tiles come from slides of different molecular subtypes. These results are consistent with the phenomenon presented in Figure 5-b and c.

## 3 Discussion

When performing a histological analysis, pathologists frequently adjust the magnification of the microscope to identify tissue features from both fine-grained and global perspectives.^57^. The presence of morphological features with substantial differences in scale adds uncertainty to the analysis of molecular traits, especially when pathologists cannot precisely define the histological or morphological biomarkers for specific subtypes. We propose a new training paradigm for WSIs - Inter-MIL to cross the chasm between subtle fine-grained features to global morphological features. By leveraging Inter-MIL, we formulate the weakly supervised learning of WSIs in the form of iterative knowledge interaction: the global slide-level features provide representative training material at the tile-level, and the optimised tile-level encoding presents a more discriminative feature space which makes the slide-level classification task easier. The results presented above demonstrate that the proposed method can train the models to successfully complete this interactive optimisation process and achieve better performance.

In our evaluation cases, most baseline methods failed to produce satisfactory subtyping results. The small number of training cases is certainly a contributing factor. This reflects the typical real-world scenario since the ground truth for subtyping based on expensive molecular profiling is often difficult to obtain on large scales. Nevertheless, with self-interactive optimisation of the tile-level embedding our Inter-MIL still achieves considerable accuracy. It should be noted that similar tasks on larger fine-curated cohorts usually require more than or close to a thousand training samples^35^, whereas our Inter-MIL achieves a similar performance level with training sets of around 100 slides. More importantly, Inter-MIL provides a more reliable set of fine-grained features which offers great improvements in the interpretation of the model. As visualised in Figures 3, S-9, S-10, and S-12, guided by more reliable attention on both the tissue level and cellular level, we can discard noisy observations and focus on the regions potentially more biologically informative. One can then analyse and quantify fine-grained image features in these regions for more representative phenotype profiling.

Results shown in Figures 3, 4, 5, S-9, S-10, S-11, S-12, S-13, S-14, S-15, S-16, and S-17 provide additional examples that demonstrate the effectiveness of the proposed optimisation method. The obtained fine-grained tile-level features could facilitate a more rational attention allocation in slide-level modelling. Hence the method contributes to enhancing the interpretability of the results. Moreover, an optimised attention allocation could provide better training resources for subsequent tile-level encoder optimisation. By observing the evolution of the attention map during multiple rounds of self-interactive training and comparing the attention regions generated by different methods, we found that Inter-MIL can automatically correct the attention distributions that do not focus on the biologically informative regions. This limitation arises from the fact that the baseline models with fixed-weight tile-level encoding^58,59^ are constrained in optimising visual features as only focus on coarse-grained morphological features^60^, which can be strongly affected by visual artefacts in limited training datasets, such as high-contrast colour and brightness.

However, re-encoding tile-level features enables the model to obtain slide-level embeddings that contain more fine-grained information. In consequence, this leads to a more accurate allocation of attention and further improves the next round of tile-level optimisation. Effectively, this self-interactive optimisation process establishes a positive feedback loop. This is demonstrated by visualizing the changes in the tile-level feature space before and after refining (as shown in Figures 5, S-13, S-14, S-16, and S-17). After a few rounds of self-interactive optimisation, the fine-grained features of the same subtype tend to automatically cluster in the latent space. Even if the initial attention distribution in a given slide may be incorrect, Inter-MIL can still rectify the attention assignment by leveraging more fine-grained representations learned from other slides. Interestingly, when we compare the tiles with the highest attention of each slide extracted by our method and to those obtained with GatedAttPool, we find that GatedAttPool provides visually mixed results with more tiles in areas marked by pen or contaminated areas (but also with higher saturation), indicating that suboptimal tiles encoding may lead to the incorrect selection of “representative tiles”. Furthermore, the examples of querying informative tile features from the feature space suggest that the WSI-based molecular subtyping task may benefit from the well-separated latent space features obtained from our method.

In a weakly-supervised setting, Inter-MIL can provide the pseudo tile-level classification prediction scores, which adds another interpretable visualization indicator beyond attention. Conventionally, assigning subtype prediction scores to tile-level patches requires fine-grained annotations^5,61^ or ROI indications^8,62^. It could be argued that one could leverage attention scores to gain insights into tile-level labels; however, these scores typically serve as indicators of importance rather than accurately translating to class prediction, especially for multi-class problems. In other words, the partial coincidence between the attention map and the prediction map for positive/negative subtypes (e.g. EMT-high/low, KRAS-yes/no, etc.) does not suggest that areas with high attention correspond to regions that respond to a specific subtype but instead represent areas that may be useful in distinguishing subtypes. Therefore, solely relying on attention regions and considering them as potential markers for specific subtypes may not be rigorous^63^. Tile-level prediction is a key contribution of our approach which greatly improves the interpretability of the regions identified by Inter-MIL. The prediction scores allow us to discard tiles without clear subtype predictions. Regions of high attention facilitate the identification of morphological correlates of molecular subtypes. Bridging between fine-grained attention at the nuclear level and the morphological features that promote molecular subtyping is the core contribution of this paper. This enables the modelling of biological evidence within the tumour microenvironment to be dynamically optimised and used to support molecular subtyping.

In addition to the Inter-MIL framework, we propose two side modules: 1) adversarial optimisation^56^ for low-attention tiles, and 2) contrastive-based pre-training^64,65^ for bag-of-tiles aggregators. The first side module encourages the model to predict ambiguously on non-important tiles, thus preventing the model from focusing on noisy regions. From the results presented in the previous section (Figures S-9, S-10), we can see that the adversarial optimisation module makes the attention area more focused. This approach also removes attention in some less critical areas. The second side module provides pretrained parameters to MIL’s aggregators, which improves the ability of the aggregators to discriminate bags of tiles in the first round of training. Although this module speeds up the convergence of the aggregator and improves the classification performance, the attention map fed back is highly sparse. The reason is that we randomly sample only a subset of the tiles in the training slides to generate contrasting bag-of-tiles and train the encoder to determine whether they are from slides of the same subtype. This encourages the model to use as few tiles as possible in each slide to distinguish subtypes. However, we have confidence in the potential of the aggregator pre-training module to improve downstream WSI analysis tasks. It’s promising if we conduct the aggregator pre-training on external large cohorts with fundamental tasks pre-training, such as tumour/benign classification^66^, or based on unsupervised pre-training like contrastive learning^64^.

In summary, we propose a novel weakly supervised MIL method for molecular subtype prediction based on histological WSIs that leverages a self-interactive algorithm to establish a connection between multi-scale features of histopathological images, allowing the efficient learning of more discriminative latent space features. Our proposed Inter-MIL framework is highly scalable and versatile, applicable not only to molecular subtyping but also can be used in typical histopathological image classification scenarios such as tumour classification^67^, prognosis^68,69^, and therapy response prediction^69^. Additionally, Inter-MIL does not require any pixel-level annotations to provide a tile-level feature pool that is biologically relevant to specific tasks, offering informative material for correlation analysis in multi-task and multi-modal scenarios^4,5,70,71^.

One limitation of Inter-MIL is that it tends to be sensitive to certain hyperparameters, such as the number of high-attention tiles selected from each slide for retraining the tile-level encoder. As shown in Figure S-7, we compared multiple values of the selected tile numbers: too many tiles lead the encoder to learn excessive noise, while too few tiles result in insufficient learning of detailed features. However, the model still outperforms the baseline under different settings of the most critical hyperparameters. A more detailed explanation of hyperparameter settings is included in the Method 4 section. Moreover, Inter-MIL relies on the first round of MIL to learn an initial attention distribution for the slides. If the first round of MIL fails or finishes too early, it may cause irreversible errors in subsequent self-interactive learning. Future work will focus on overcoming these technical challenges, validating the framework’s performance in a broader range of tasks, and exploring the reusability value of the generated tile-level feature pool in other tasks. In addition, we plan to explore the conjunction of Inter-MIL with other network architectures such as Graph Networks^21^ or Vision Transformers^23^, which explicitly capture contextual information spatial-awarely.

## 4 Method

### TCGA cohorts preparation

In this section, we describe in detail the preparation of experimental datasets (mentioned in Section 2) used in this study. The slide sets and metadata are collected from the TCGA repository^55^. We use only the digitized Formalin-Fixed Paraffin-Embedded (FFPE) slides, as it is the gold standard for histopathological diagnosis. We exclude the following cases due to technical artefacts: 1. In TCGA-OV cohort, we exclude 37 WSIs without available EMT-score; 2. For TCGA-COAD cohort, we exclude 413 WSIs out of 459 and for TCGA-LUAD cohort, we exclude 475 WSIs out of 541 as their labels on KRAS status are labelled as ‘Not Available’ or ‘Unknown’. The remaining 46 WSIs from TCGA-COAD and 66 WSIs from TCGA-LUAD are combined as the experimental dataset for task *COLU-KRAS*; 3. In TCGA-LUAD cohort, we exclude 280 WSIs out of 541 with labels on EGFR as ‘Not Available’ or ‘Unknown’; 4. In TCGA-BRCA cohort, we exclude 718 WSIs out of 1133 with labels on HER2 as ‘Not Evaluated’.

For each slide, we extract 256 *×* 256 tiles without overlap at 40 *×* magnification (0.25*μm* per pixel) from tissue regions. These morphomolecular subtyping tasks are challenging not only because of the nature of the problem but also because of the small cohort size as well as a combination of different tissue types (*COLU-KRAS*), and severe class imbalance (*LU-EGFR* and *BR-HER2*). To ensure enough training samples, we split the data into training/test sets with the following ratio: 70% / 30% for *OV-EMT* and *COLU-KRAS*, and 50% / 50% for *LU-EGFR* and *BR-HER2*.

Next we comment on our annotation protocol. In the *OV-EMT* task, the continuous EMT scores, which range between 0 and 1, are derived from the results of sequencing analyses conducted by our collaborating pathologists. As per the pathologists’ assessment, the median EMT score of the entire cohort is used to distinguish between EMT-low/high. For the other three tasks, all annotations can be found in the clinical records of TCGA, and they are discrete labels: ‘YES/NO’ for KRAS and EGFR status in TCGA-COAD and TCGA-LUAD, and ‘Positive/Negative’ for HER2 status in TCGA-BRCA.

### Whole slide images preprocessing

The preprocessing of WSIs involves the following steps: 1) Removing grey or white background; 2) Removing blue and green contaminated areas; 3) Removing markers of the red, green and blue pens. All these preprocessing steps are conducted with OpenCV tools (pypi.org/project/opencv-python/). Then, we divide WSIs into tiles, and we discard the tiles with a proportion of tissue less than 70%.

### Deep learning model implementation

The source code and detailed environment configuration files are available at github.com/superhy/LCSB-MIL. The deep learning model is implemented using the PyTorch-1.6 framework. Training and testing of our model were performed on four NVIDIA RTX 2080Ti GPUs.

The following settings are used in all tasks unless specified otherwise. For the two modules of MIL workflow, we use ResNet-18 pretrained on ImageNet^25^ as the vision encoder (encoder) to generate the initial feature embedding of tiles, and we apply the Gated-AttPool^28^ as the instances-bag aggregator (aggregator) and the subtyping classifier, which contains three fully connected layers for classification and one attention-based pooling layer. Furthermore, we choose the weighted cross-entropy (WCE) loss to tackle the class imbalance. The Adam optimisation with a learning rate of 0.0001 is applied, and we set the batch size of 8 WSIs for training the aggregator and 128 tiles for the encoder. In the training stage, each interactive round consists of 5 epochs for the aggregator and 2 epochs of vision encoder training. We set a delayed stop mechanism in the aggregator training of the first round to improve the initial stability of the proposed Inter-MIL framework. Moreover, we set the convergence point at which the overall training of Inter-MIL stops, and we describe these parameters in detail in the following paragraphs.

### Overall framework of Inter-MIL

The proposed Inter-MIL framework consists of two learnable neural network modules: Module-1, the instance-bag aggregator (aggregator) based on Gated Attention Pooling network, and Module-2, the trainable tile-level feature encoder based on ResNet-18. These modules are optimised alternatively until convergence conditions are met, as illustrated by Figure 6, which depicts the details in the main framework of Inter-MIL method as well as supporting modules.

**Figure 6.**
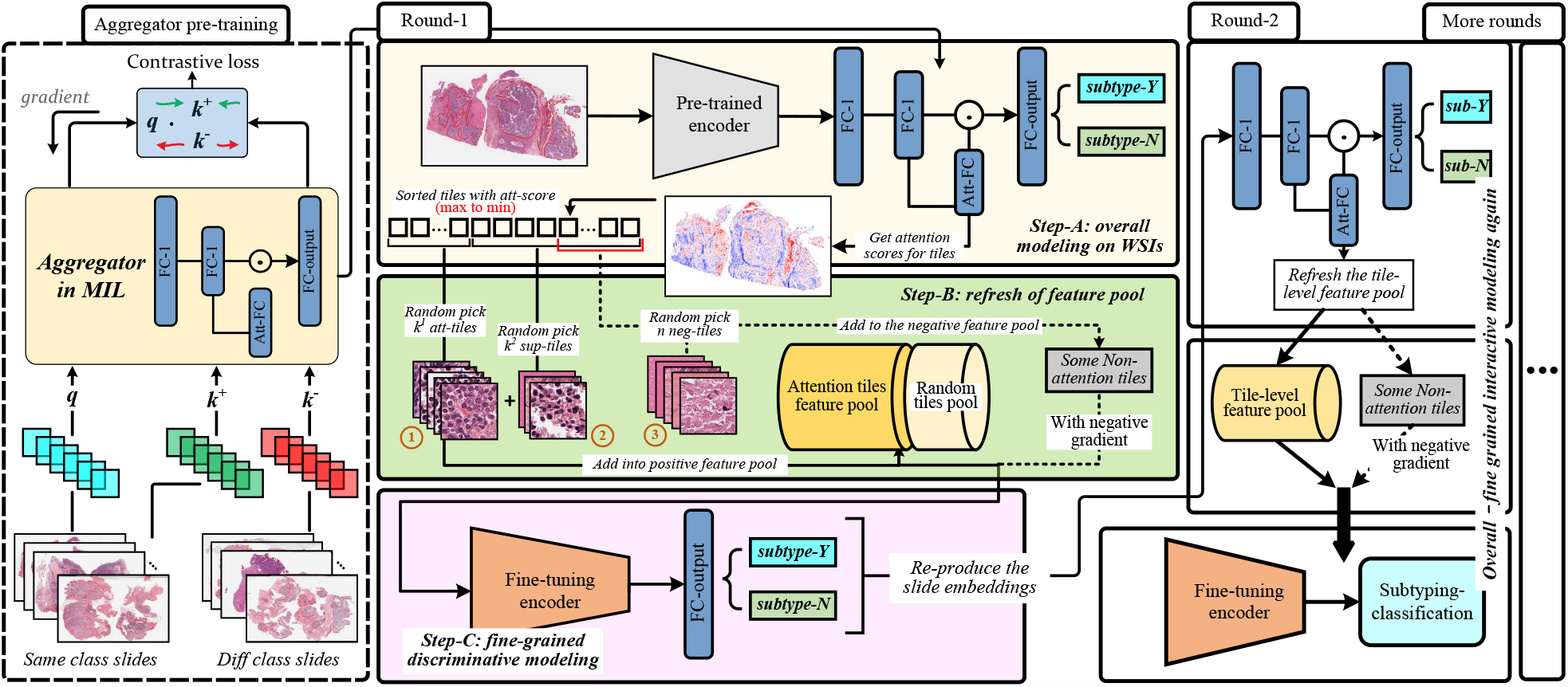
Overview of the Inter-MIL framework. The framework is divided into three parts, from left to right, highlighting its various components and functions. **Left**, The aggregator is pretrained using contrastive learning, where embeddings of slides of the same and different subtypes are fed into the aggregator in pairs. The training objective is to minimize the distance between embeddings from the same subtype while maximizing the distance between embeddings from different subtypes. **Middle**, The self-interaction MIL algorithm consists of three steps within each round: 1) train AttPool network with pretrained tile embeddings to obtain the attention value for each tile, 2) constructing a tile-level feature pool with high-attention tiles and supplementary tiles (defined in Eq. 4), optionally including low-attention tiles (defined in Eq. 5), and 3) fine-tuning the CNN encoder (using ResNet) with the tile-level feature pool. **Right**, The tile embeddings are reproduced for the next round of AttPool training, and the subsequent rounds of self-interaction MIL continue until convergence is achieved.

To begin with, given a WSI with *L* tiles 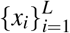 and the tile-level feature encoder *f*_*cnn*_(·), we let 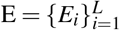 be the set of tile embeddings, such that *E*_*i*_ = *f*_*cnn*_ (*x*_*i*_). The dimension of tile embedding is *E*_*i*_ ∈ ℝ^512^ . In Module-1, the AttPool or Gated-AttPool based aggregator^20,28^ receives tile embeddings *E* and outputs a classification result *y*_*cls*_:

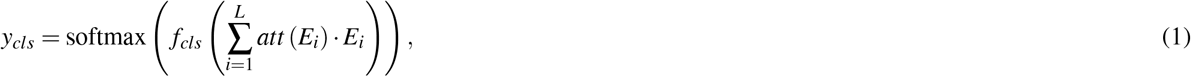

where *f*_*cls*_(·) is the output layer for classification, the attention score *att*(*E*_*i*_) ∈ [0, 1] reflects the contribution of the *i*-th tile to the classification, and 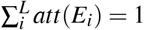. We train aggregator by optimising the slide-level prediction, which can be formalized as follows:

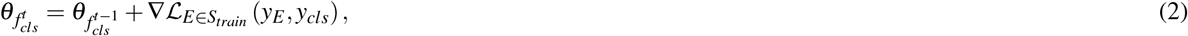

where *t* and *t* − 1 indicate the current and previous training loops, the tile embedding set *E* comes from training slide set *S*_*train*_, and *y*_*E*_ denotes its subtype label.

Here, Inter-MIL aims to train fine-grained histological features on representative tiles in multiple rounds, which helps to optimise the WSI embedding set *E* for the aggregator. In Module-2, to refresh tile embeddings *E*^*t*−1^ to *E*^*t*^ for the *t*^*th*^ training loop, we use *k* representative tiles 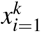 with high attention scores to fine-tune 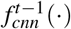 to 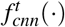, which encodes the fine-grained features. The training of 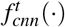 aims to optimise prediction loss at the tile level. After this step, we regenerate the tile embeddings *E*^*t*^ using the fine-tuned feature encoder 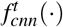 and continue with the next training round for the aggregator:

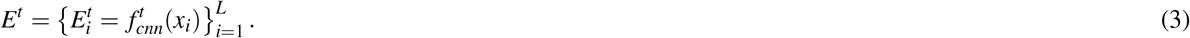

As shown by Figure 6, in the middle column, the optimisation of Module-1 (beige block) and Module-2 (pink block) constitutes a training loop (round) of Inter-MIL, while the right column represents subsequent training rounds, which repeats the switching and interaction in overall and fine-grained feature optimisation until convergence. Constructing tile-level training materials (shown in green blocks) is a critical step in the Inter-MIL framework.

We now elaborate on the strategy for selecting representative training tiles. Our target is to construct a tile-level feature pool *S*^*pos*^. Given a WSI, we rank tiles based on their attention scores in a monotonically decreasing order, i.e. 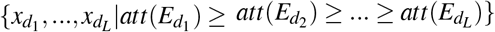, where *att*(*E*_*i*_) refers to Eq. 1. We define a set of **attention tiles** *S*^*top*^ as a set of randomly sampled *k*^1^ tiles out of the top *K* highest attention tiles, i.e. 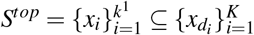. Similarly, we define a set of **supplementary tiles** *S*^*sup*^ as a set of randomly sampled *k*^2^ tiles out of the remaining *L*−*K* tiles, i.e. 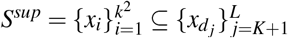. *K* is determined by the total number of tiles *L* separately for each WSI. *S*^*sup*^ increases the diversity of tile features that may not be captured by *S*^*top*^. We construct the set *S*^*pos*^ of tile-level feature pool as follows:

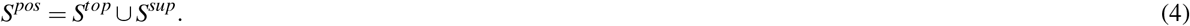

The main goal of training on the representative tile feature set *S*^*pos*^ is to optimise the representation of tile-level feature encoders for fine-grained histological features. However, to attenuate the influence of noisy tile-level features. We also want the encoder to learn to distinguish and discard non-relevant tiles that could end up being allocated high attention scores. Therefore, we construct a set of **negative tiles** *S*^*neg*^ as:

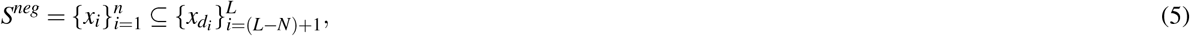

which consists of *n* randomly sampled tiles from the *N* lowest attention tiles from each WSI. We adopt a different optimisation strategy for negative tiles *S*^*neg*^ than for attention tiles *S*^*top*^ and supplementary tiles *S*^*sup*^. More details on the optimisation strategy for *S*^*neg*^ are provided in the next subsection. The pseudocode for constructing the tile-level feature training repository is provided in Algorithm 1.

### Optimisation of tile-level feature encoder

As with standard MIL training, the aggregator is optimised based on the slide-level annotation *y*_*s*_ of slide *s* and the feed forward process shown in Eq. 1. The aggregator is optimised according to loss of weighted cross-entropy: ℒ (*y*_*s*_, *y*_*cls*_). Additionally, the encoder *f*_*cnn*_(·) is trained based on the tile-level feature pool *S*^*pos*^ (and *S*^*neg*^) described in the previous section. The optimisation process is as follows:

#### Algorithm 1

Construction of tile-level fine-grained feature pool (for each slide)

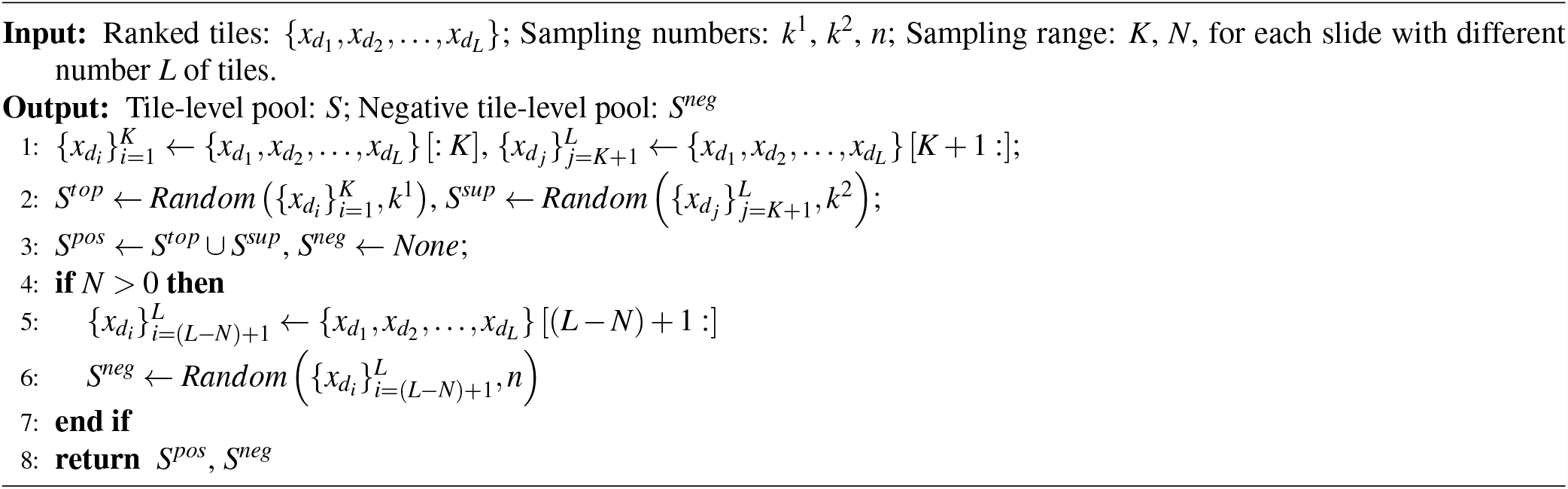

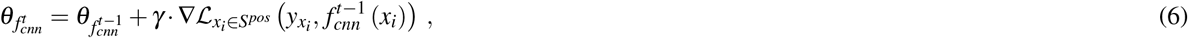

where 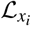 refers to the loss of tile-level inputs, *θ* denotes the weights of the encoder, and *γ* is the learning rate. 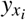 is the annotation of tile *x*_*i*_ inherited from the slide it belongs to (i.e., *y*_*s*_).

In contrast to modelling representative tile-level information in *S*^*pos*^, we aim to reduce noise interference in the model. Treating low-attention tiles in *S*^*neg*^ as noise, we train the model to discard these tile-level features using an adversarial optimisation approach^56^, This approach trains the encoder to classify the noisy samples as poorly as possible by back propagating the negative gradients. Thus, we extend the optimisation of the encoder *f*_*cnn*_(·) in (6) to incorporate *S*^*neg*^ as follows:

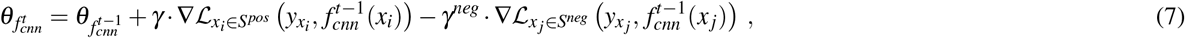

where *γ*^*neg*^ denotes the learning rate of the negative training.

The overall optimisation and prediction pseudocode of InterMIL is given by Algorithm 2.

### Contrastive pre-training of instance-bag aggregator

We designed an additional module to improve the convergence of the instance-bag aggregator (slide-level classifier) and bestow the aggregator with a better initialization that increases the training stability.

Inspired by self-supervised contrastive learning algorithms^64,65^, we transplant the contrastive learning based on sample data enhancement to the scenario of WSI classification. Thus we can pre-train the MIL model for the recognition ability of tile-level bags. Unlike unsupervised contrastive learning, which determines whether augmented samples are of the same origin, we currently apply supervised annotations to evaluate whether randomly sampled tile bags belong to the same subtype. Given 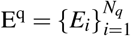 as a query bag of tile embeddings randomly picked from slides of any subtype, where *N*_*q*_ is the sampling quantity, 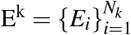 is the key embedding bag from slides of the same or different subtype with E^q^, where *N*_*k*_ = *N*_*q*_. The learning target of the aggregator pre-training is to reduce the distance between tile embedding bags from the same subtype, while increasing the distance between tile embedding bags from different subtypes. This results in the following loss function:

#### Algorithm 2

Histopathology subtyping workflow of Inter-MIL framework

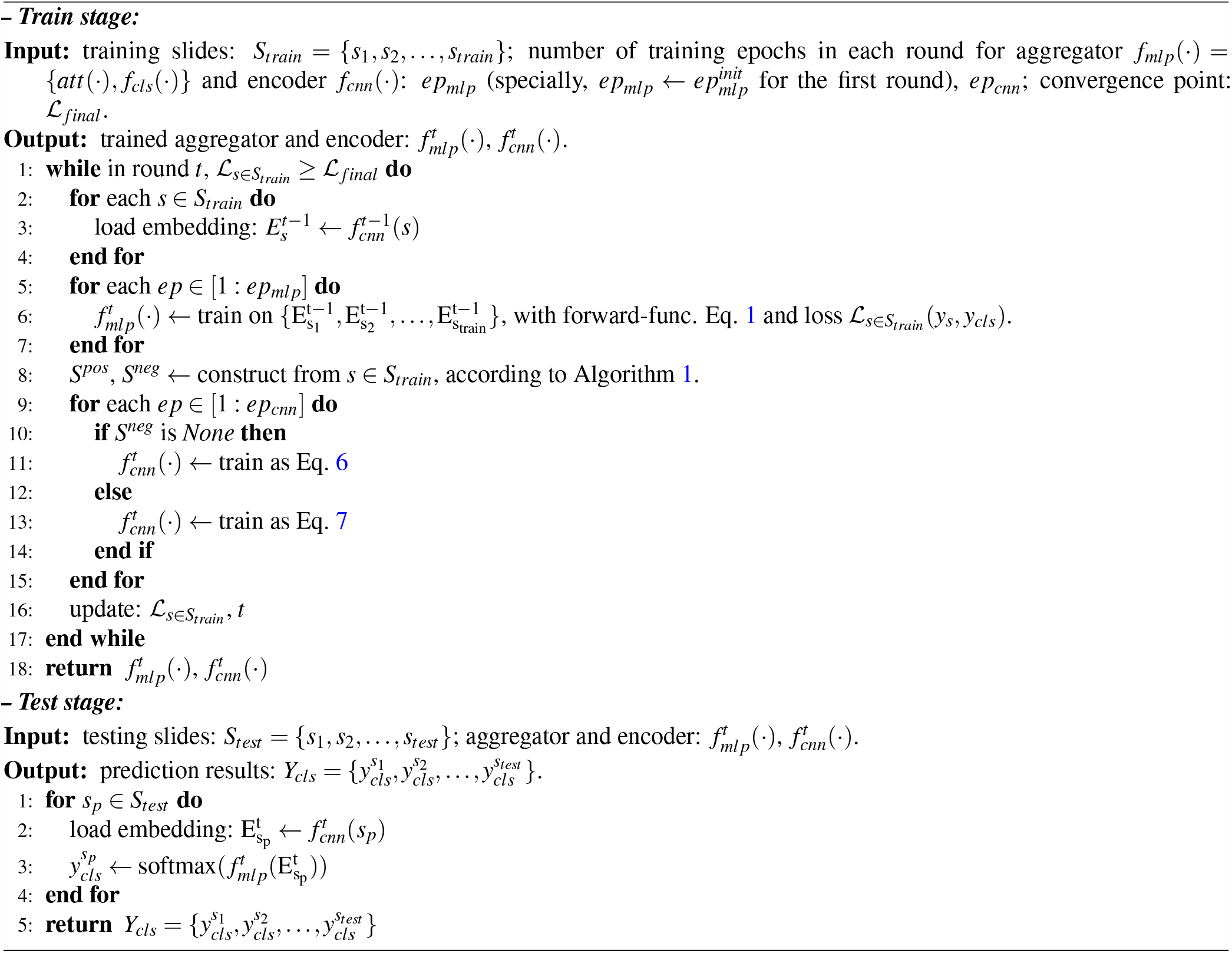

#### Algorithm 3

Contrastive training for instance-bag aggregator

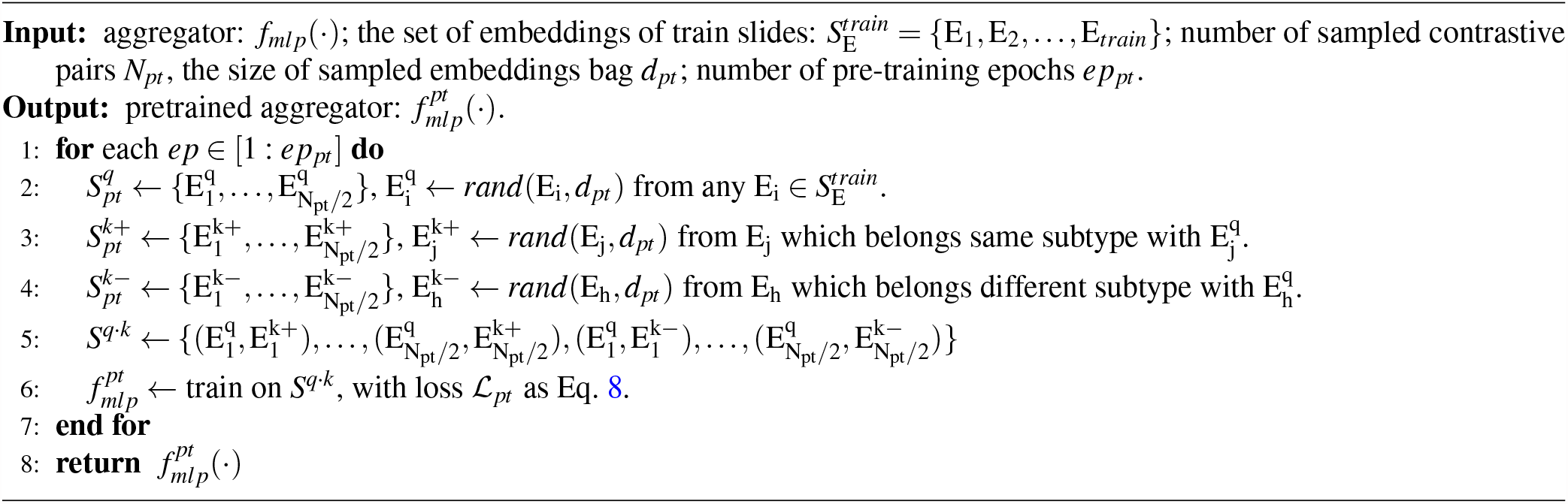

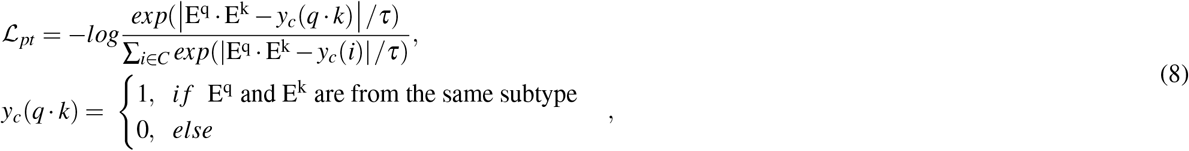

where *y*_*c*_(·) ∈ {0,1} indicates if E^q^ and E^k^ are from the same subtype and *C* = 2, *τ* is a sensitivity parameter and we set *τ* = 1.

The leftmost part of Figure 6 illustrates a schematic of the optional module for aggregator pre-training, and Algorithm 3 presents its pseudocode.

### Hyper-parameters in Inter-MIL

In the experiments, we set the hyperparameters for Inter-MIL as listed in Table 3. We adopt a delayed stop strategy for training the aggregator in the initial round to ensure that valuable high-attention tiles are selected. Specifically, 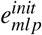 will continue to train until the loss value drops to *ℒ*_*init*_. The number of epochs for aggregator pre-training uses *v*_1_ and *v*_2_ based on the size of the training cohorts. For tasks *OV-EMT* and *COLU-KRAS*, due to the smaller training set, we opt for *v*_1_, while for tasks *LU-EGFR* and *BR-HER2*, with the larger training sets available, we chose *v*_2_ to enable longer pre-training.

**Table 3.** Setting of hyperparameters.

| Parameter | Value | Notation |
| --- | --- | --- |
| $K$ | $0.05 \times L$ | Selection range of high-attention tiles, for fine-tuning tile-level encoder. |
| $N$ | $0.2 \times L$ | Selection range of low-attention tiles, for denoising with adversarial optimisation. |
| $k^1$ | 50 | The number of attention tiles for fine-tuning tile-level encoder. |
| $k^2$ | $0.4 \times k^1$ | The number of supplementary tiles for fine-tuning tile-level encoder. |
| $n$ | $0.2 \times k^1$ | The number of negative tiles for denoising on the tile-level encoder. |
| $ep_{mlp}$ | 10 | In each round of interaction, the training epoch of aggregator. |
| $ep_{mlp}^{init}$ | Dynamic | In the initial round of interaction, $ep_{mlp} \leftarrow ep_{mlp}^{init}$, the end of $ep_{mlp}^{init}$ depends on $\mathcal{L}_{init}$. |
| $ep_{cnn}$ | 2 | In each round of interaction, the training epoch of tile-level encoder. |
| $N_{pt}$ | 6000 | The number of sampled contrastive pairs, $N_{pt} = N_{k+} + N_{k-}$, and $N_{k+} = N_{k-} = N_q$. |
| $d_{pt}$ | 8000 | The size of the bag of tiles sampled from each slide. |
| $ep_{pt}$ | $v_1 = 30, v_2 = 50$ | The number of epochs for aggregator pre-training, for tasks of *OV-EMT* and *COLU-KRAS*, pick $v_1$, for tasks of *LU-EGFR* and *BR-HER2*, pick $v_2$. |

### Evaluation setting up

We compare our proposed Inter-MIL method with several state-of-the-art MIL algorithms, including CNN-MIL^9^, AttPool^20^, Gated-AttPool^20,28^, CLAM^13^, and FocAtt-MIL^14^. To provide a direct comparison with our method, we select Gated-AttPool with a fixed pretrained CNN encoder as the baseline. We randomly select training sets without replacement for 10 folds in the *OV-EMT* task and 5 folds for the other tasks. For each evaluation fold, we use the remaining samples, excluding those in the training set, as the test set. Due to the limited amount of training data in *OV-EMT* and *COLU-KRAS* tasks, we do not split the data further into a validation set. In all experiments, we use the last epoch’s model for testing. We use the output of the aggregator as the final prediction of the Inter-MIL. We empirically determine the value of ℒ *final* based on the performance of the baseline model, and set ℒ_*init*_ as ℒ_*final*_ + 0.15. We record the performance in terms of the average and variance of the area under the receiver operating characteristic curve (AUC) with macro averaging across multiple test folds.

## Data Availability

All data produced in the present study are available upon reasonable request to the authors.

## Acknowledgements

This work was supported by: Y.H., C.V. and J.R. - National Institute for Health Research (NIHR) Oxford Biomedical Research Centre; K.S., C.V., and J.R. - Innovate UK funded PathLAKE consortium; K.G. - Clinical Lectureship from the National Institute for Health Research (NIHR, grant no. CL-2017–13-001); R.W. - EPSRC Centre for Doctoral Training in Health Data Science (EP/S02428X/1) and Oxford CRUK Centre for Cancer Research. Computation used the Oxford Biomedical Research Computing (BMRC) facility.

We thank Professor Ian Mills and Oxford Prostate Cancer Biology Group for their guidance and suggestions on molecular biology for this paper.

## Author contributions

Y.H., K.S., and J.R. conceived the study. Y.H. conducted the experiments. K.G., M.W., D.W., and C.V. checked the correctness of biology and pathology. K.G. prepared a part of the annotations. Y.H. and K.S. wrote the paper. B.L., W.B., M.W., and R.W. did proofreading and edited the manuscript. B.L. rigorously checked the technical details and description of the results. W.B. validated some of the follow-up interpretability results. N.K.A. developed and maintained visualization and data annotation tools. S.M. contributed some code and is responsible for server maintenance. A.A. led the research project in the biomedical section. C.V. and J.R. led the research project in the computational section. All authors reviewed and edited this paper.

## Supplementary information

**Figure S-7.**
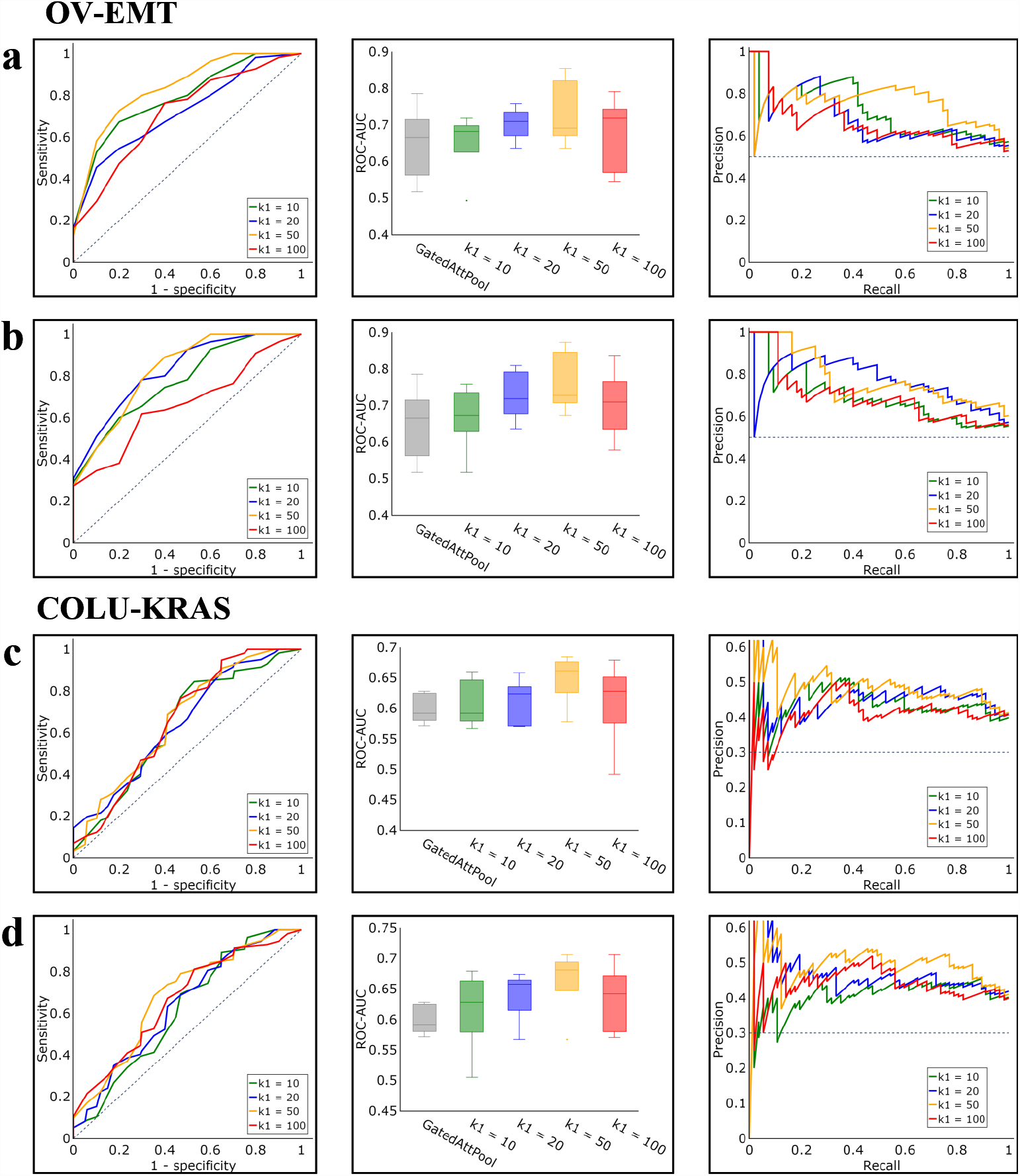
Fluctuations in performance with various values of parameter *k*^1^, which could be 10, 20, 50 (used), and 100. **a** and **b** refer to the methods of Inter-MIL and adInter-MIL, on the task of *OV-EMT*, while **c** and **d** show the performance of Inter-MIL and adInter-MIL, on the task of *COLU-KRAS*. For **a, b, c**, and **d**, the left shows the AUC-ROC curve under different values of *k*^1^, the mid shows the result comparison of ROC-AUC, and the right shows the Precision-Recall Curve (PRC) under different values of *k*^1^.

**Figure S-8.**
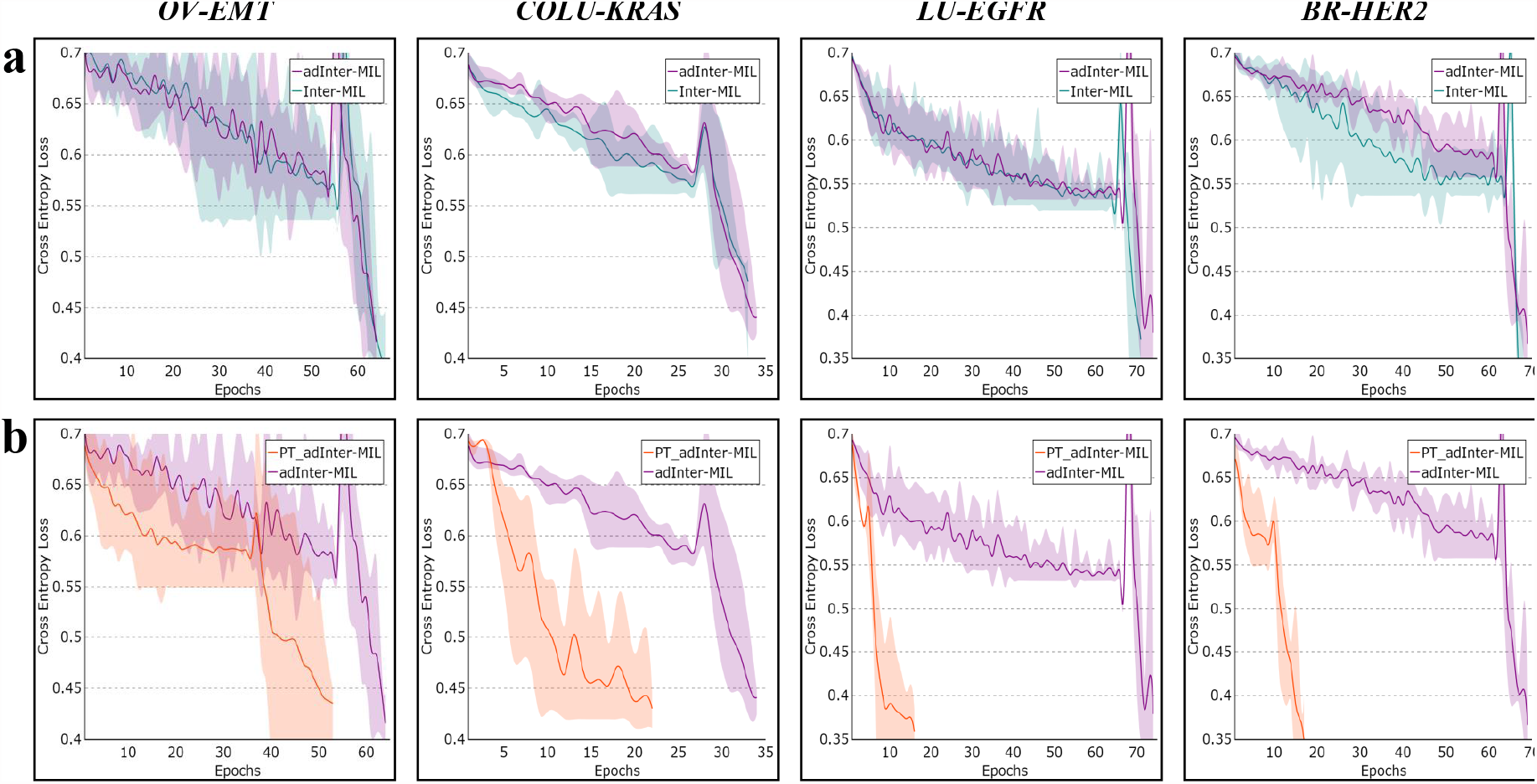
More comparison on the log of training loss. **a**, the training logs of Inter-MIL vs adInter-MIL. **b**, the training logs of adInter-MIL vs PT-adInter-MIL. From left to right, for tasks *OV-EMT, COLU-KRAS, LU-EGFR*, and *BR-HER2*.

**Figure S-9.**
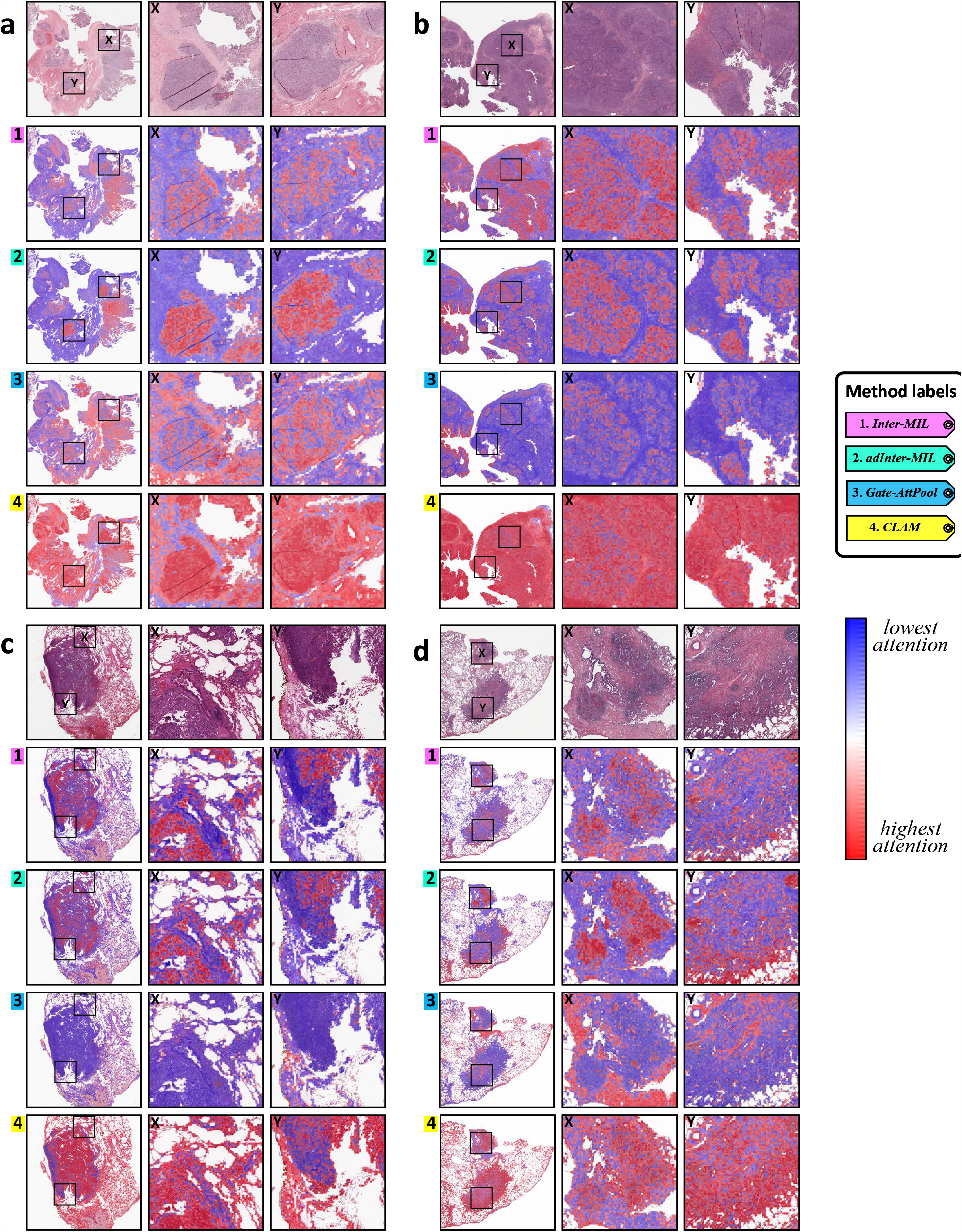
Interpretable attention heatmaps produced by different methods. **a - d**, Respectively, an EMT-low case, an EMT-high case, a KRAS-no case, and a KRAS-yes case.

**Figure S-10.**
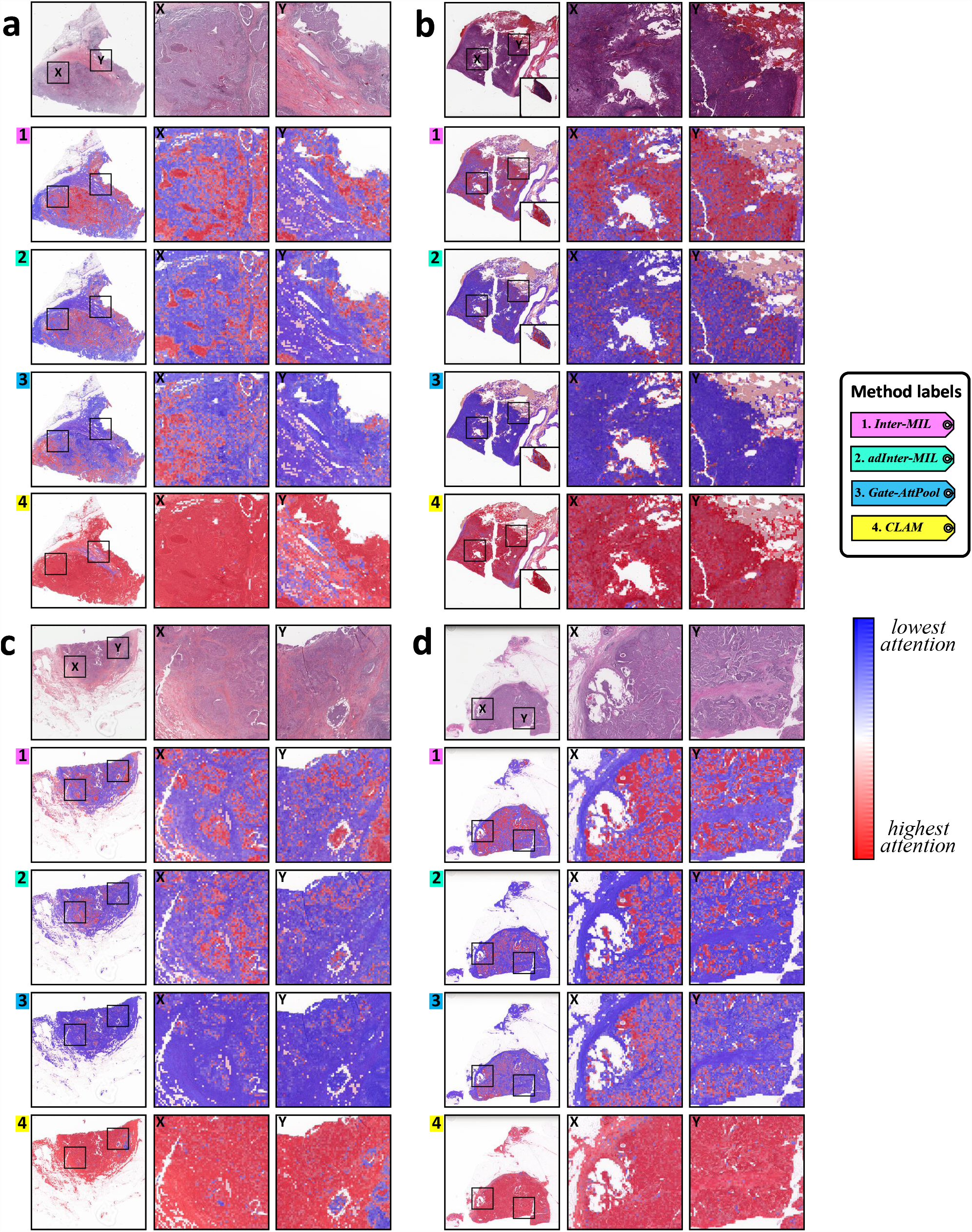
Interpretable attention heatmaps produced by different methods. **a - d**, Respectively, an EGFR-no case, an EGFR-yes case, a HER2-negative case, and a HER2-positive case.

**Figure S-11.**
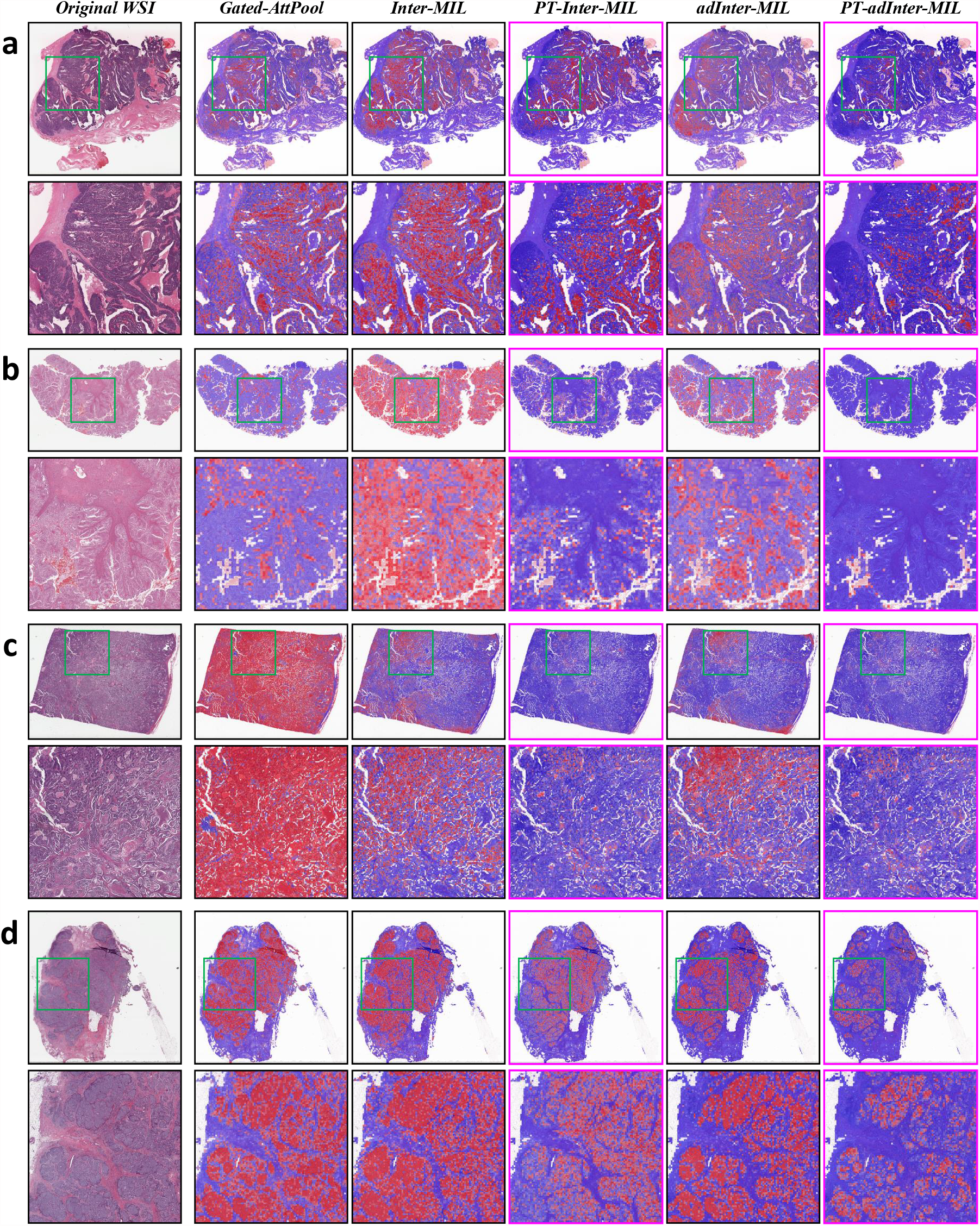
Comparison of attention heatmaps produced by methods with/without the pre-training module for aggregator. **a - d**, The demo case from tasks of *OV-EMT, COLU-KRAS, LU-EGFR*, and *BR-HER2* respectively.

**Figure S-12.**
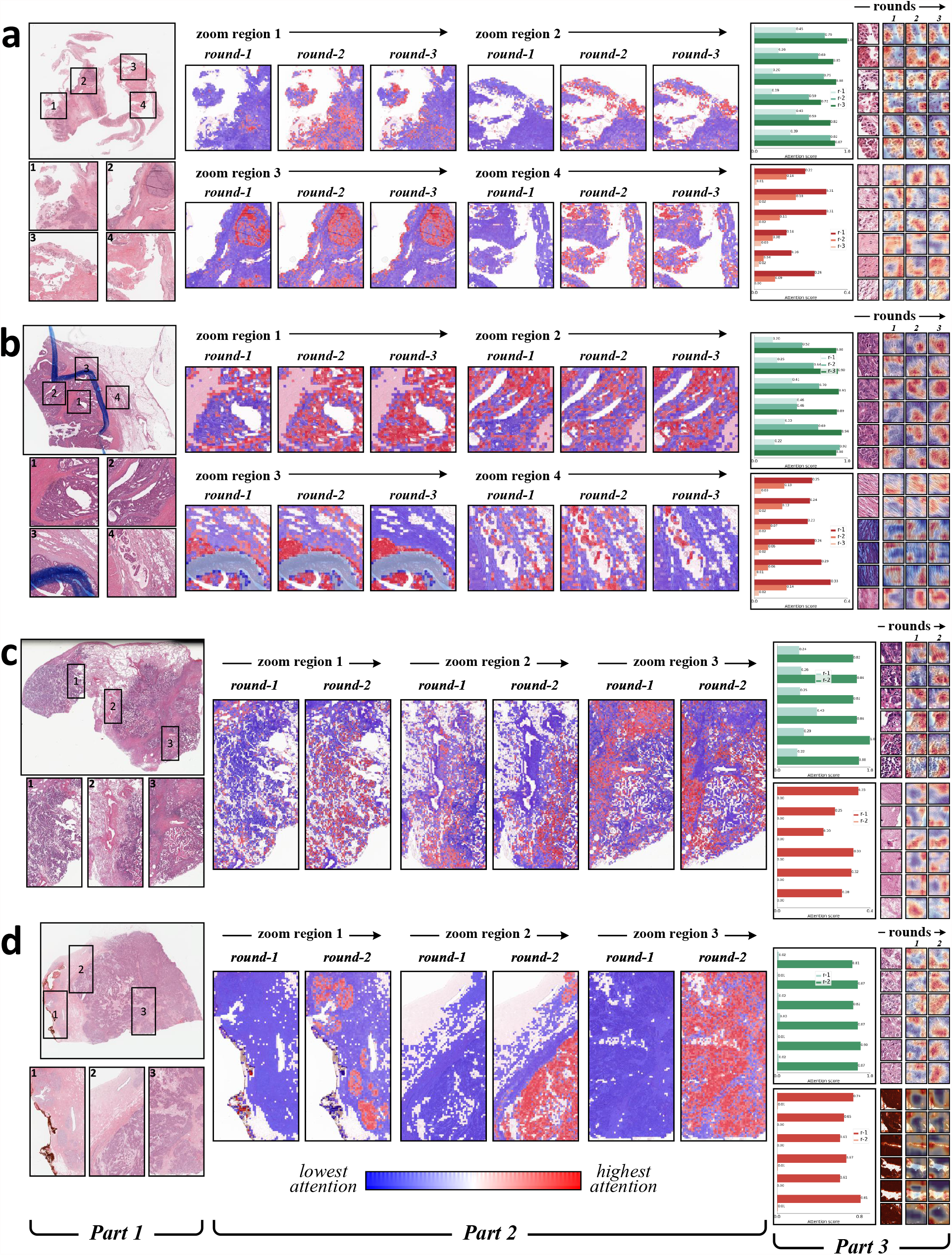
More demos of tile-level features’ attention evolution. **a - d**, The demo case from tasks of *OV-EMT, COLU-KRAS, LU-EGFR*, and *BR-HER2* respectively. For **a - d**, from left to right, each shows the original WSI, the evolution of attention heatmap in some highlighted regions, and then the attention value evolution on some examples of high informative tiles and noise tiles accompanied with their fine-grained heatmaps.

**Figure S-13.**
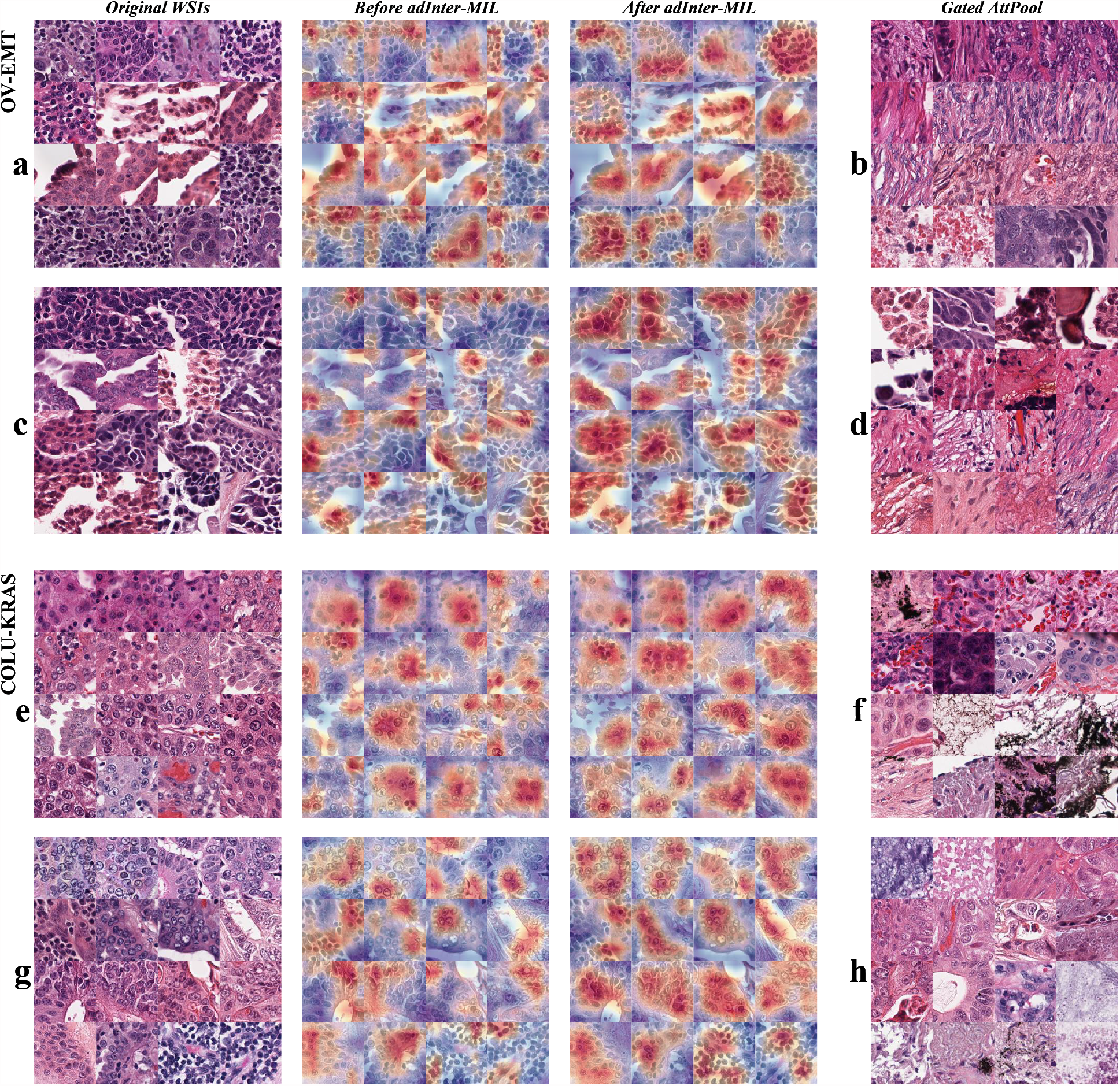
Examples of tiles with the top attention value by methods: adInter-MIL and GatedAttPool. **a, c, e, g** examples of top attention tiles from adInter-MIL. **b, d, f, h** examples of top attention tiles from GatedAttPool. For **a, b, c, d, e, f**, and **g, h**, the examples are from EMT-low/high and KRAS-no/yes cases. For **a, c, e** and **g**, the left column shows the original WSIs, the middle shows their fine-grained heatmaps before applying adInter-MIL, and the right shows their fine-grained heatmaps after applying adInter-MIL.

**Figure S-14.**
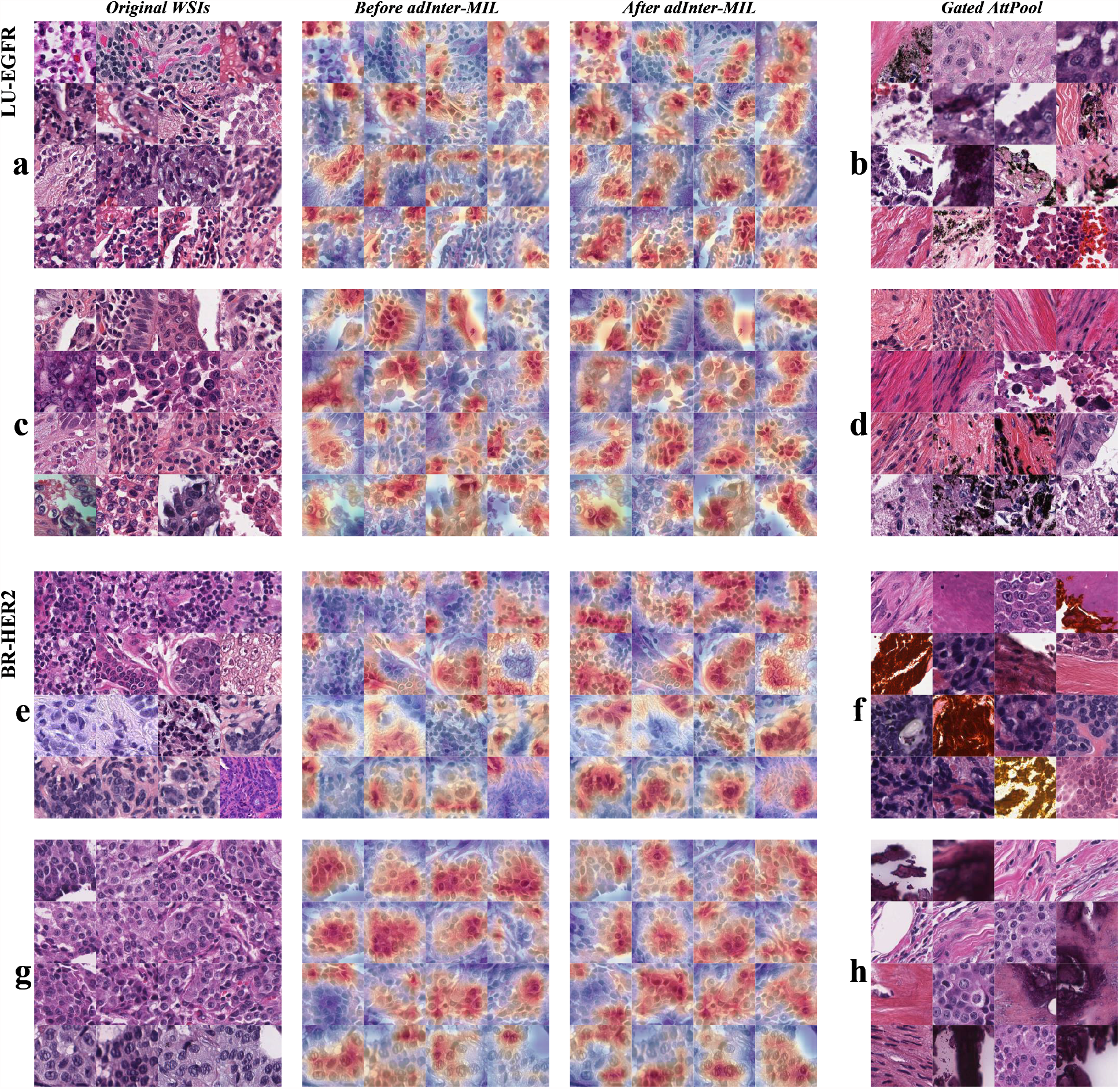
Examples of tiles with the top attention value by methods: adInter-MIL and GatedAttPool. **a, c, e, g** examples of top attention tiles from adInter-MIL. **b, d, f, h** examples of top attention tiles from GatedAttPool. For **a - b, c - d, e - f**, and **g - h**, the examples are from EGFR-no/yes and HER2-neg/pos cases. For **a, c, e** and **g**, the left column shows the original WSIs, the middle shows their fine-grained heatmaps before applying adInter-MIL, and the right shows their fine-grained heatmaps after applying adInter-MIL.

**Figure S-15.**
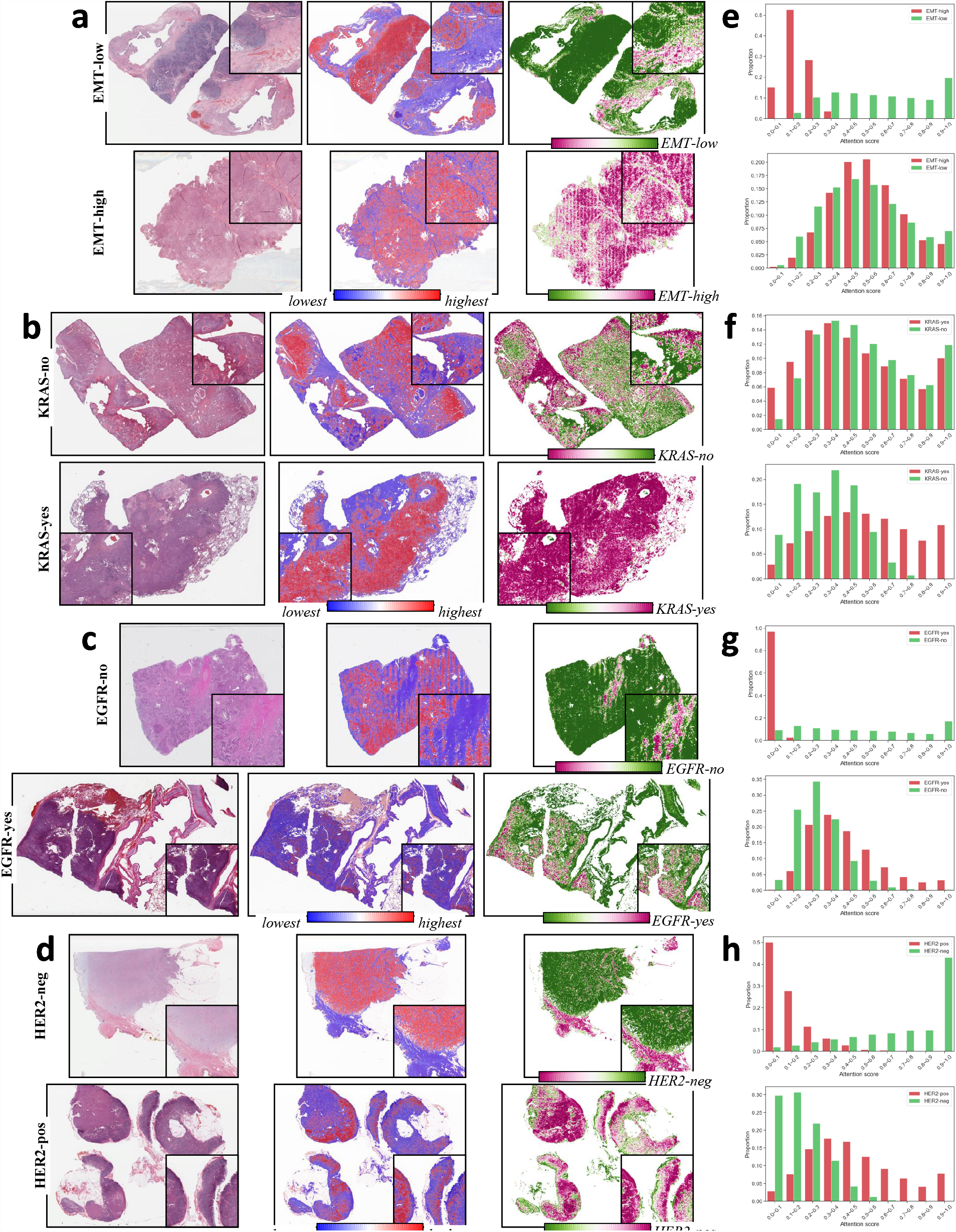
More demos on distributions of slide-level attention and classification score. **a - d**, The cases from tasks: *OV-EMT, COLU-KRAS, LU-EGFR*, and *BR-HER2*. For each, the cases from EMT-low/high, KRAS-no/yes, EGFR-no-/yes, and HER2-neg/pos are separated as top/bottom half. **e - h**, Respectively, the proportion of tiles with prediction results for EMT-low/high, KRAS-no/yes, EGFR-no/yes, and HER2-neg/pos. In which the top/bottom separation is similar to (a - d).

**Figure S-16.**
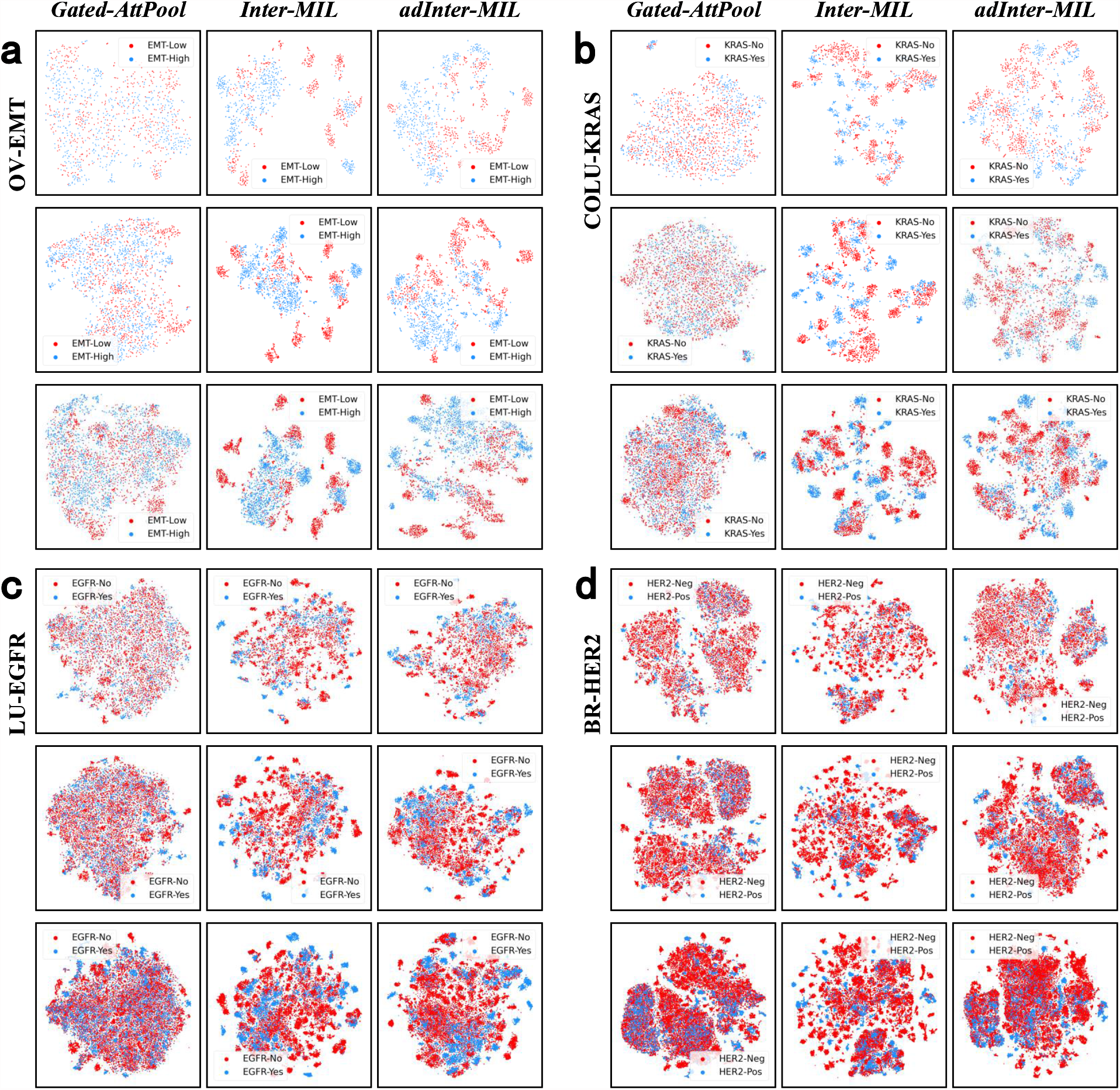
More visualization of feature representation obtained from different methods. **a - d** show the outcomes of tasks *OV-EMT, COLU-KRAS, LU-EGFR*, and *BR-HER2*. In which, the left column, middle column, and right column indicate methods GatedAttPool, Inter-MIL, and adInter-MIl respectively. In addition, the first, second and third rows of **a - d** represent the 50, 100, and 200 tiles with the highest attention as the feature samples.

**Figure S-17.**
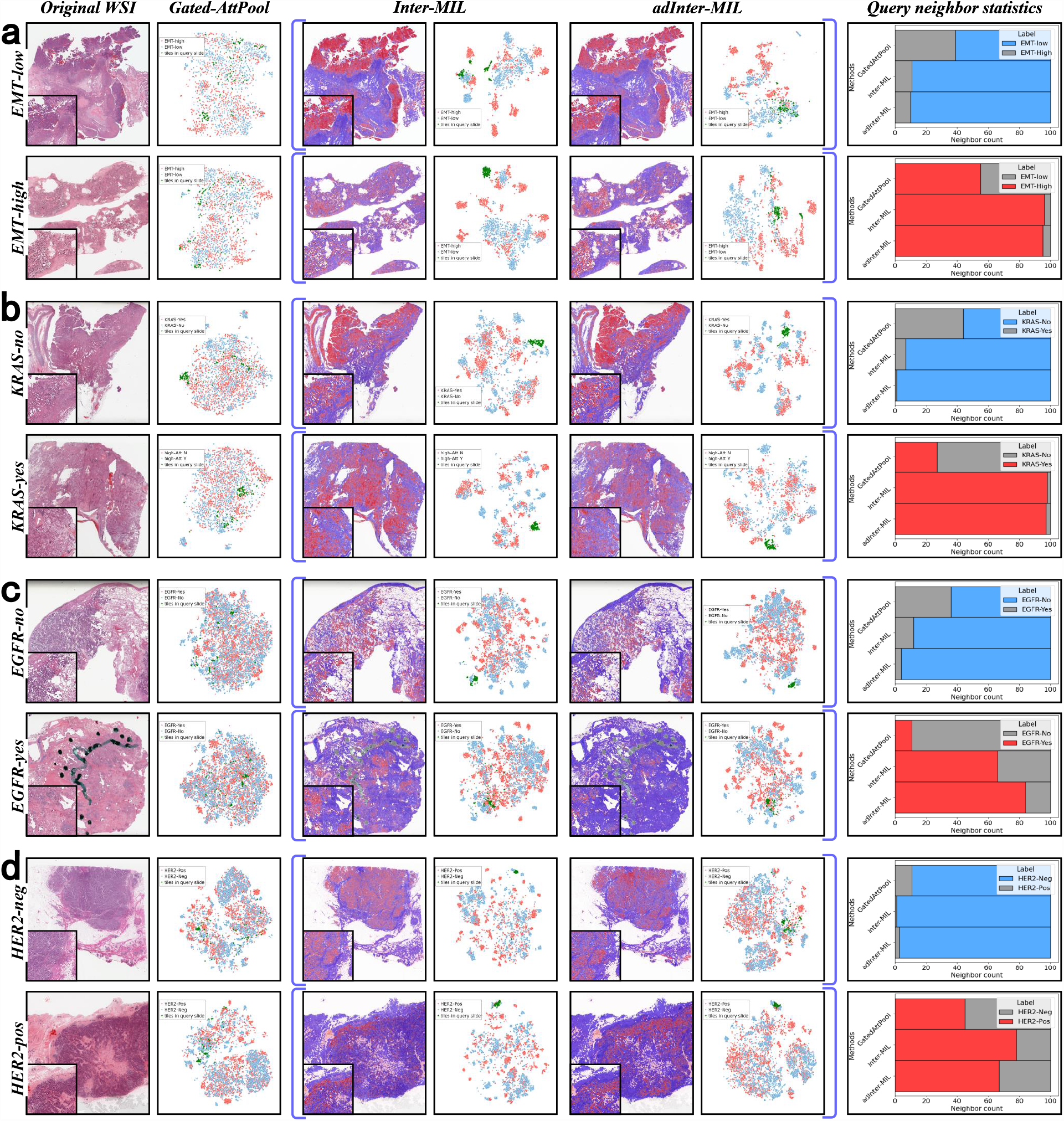
More demos of querying high-informative tiles from different feature spaces. **a - d** show the cases from tasks *OV-EMT, COLU-KRAS, LU-EGFR*, and *BR-HER2*. In **a - d**, the top row shows examples with the negative label (low, no, or negative), and the bottom row shows examples with the positive label (high, yes, or positive). For columns from left to right, respectively, shows the feature spaces of the original WSI and baseline methods, the attention heatmaps of the Inter-MIL method and adInter-MIL, and the feature spaces they generate are shown, respectively. In the feature space, green points represent the query feature distribution of example slides. The right column shows the proportion of the nearest 100 tiles when querying the demo slides in the feature space, blue refers to the negative category (low, no, or negative), and red refers to the positive class (high, yes, or positive).

